# Distinct Multimodal Brain Profiles Bridging Age of First Alcohol Use and Subsequent Substance Use in Young Adults

**DOI:** 10.64898/2026.09.30.26364406

**Authors:** Kayla Hannon, Nathaniel Harnett, You (Lily) Cheng, Julia E. Cohen-Gilbert, Cole Korponay, Poornima Kumar, Marisa M. Silveri, Lisa Nickerson

**Author notes:** **Corresponding authors:** Kayla Hannon, Lisa Nickerson. These authors contributed equally (Kayla Hannon, Nathaniel Harnett). **Disclosures:** The authors of this study have no disclosures.

## Abstract

Age of first alcohol use is associated with alterations in brain development and an increased risk of problematic substance use later in life. However, it remains unclear if neurobiological alterations associated with first-use age are present in young adulthood and how these alterations may be associated with different patterns of adult substance use. In this study, we employed SuperBigFLICA, a semi-supervised multimodal imaging data fusion approach, to identify joint structural morphometric and functional MRI (fMRI) brain profiles present in adulthood that are most predictive of age of alcohol first-use in the Human Connectome Project-Young Adult cohort (n=622). Our model achieved moderate performance (r=0.22 in external validation), identifying multimodal structure-function profiles predictive of first-use largely involving frontal regions. We then utilized Canonical Correlation Analysis to determine whether these multimodal brain profiles were differentially linked to patterns of adult substance use behaviors. We identified three significant brain modes (Mode 1: *r* = 0.2750, *p* = 0.014; Mode 2: *r* = 0.2249, *p* = 0.005; Mode 3: *r* = 0.1568, *p* = 0.029) associated with distinct profiles of alcohol and cannabis use. Together, we identified multimodal brain patterns in young adults that predict age of first use of alcohol and demonstrated that resulting multimodal structure-function brain profiles capture distinct multivariate patterns of adult substance use, providing a more integrated view of substance use neurobiology.

## Introduction

An estimated 70.7% of individuals in the United States report first using alcohol before the age of 21 (Substance Abuse and Mental Health Services Administration, 2025), with the peak rates of initiation occurring between 7^th^ and 11^th^ grade (Johnston et al., 2022). Earlier age of first alcohol use (AFAU) is associated with heavier alcohol consumption, increased binge drinking, and greater risk of alcohol and polysubstance use disorder later in life (Brennan et al., 2025; Flores-López et al., 2026), with risk increasing with younger AFAU (Gardner et al., 2024).

Understanding the neurobiological correlates of AFAU may help identify developmental pathways with later substance use vulnerability. The frontal cortex has emerged as a key region showing structural and functional differences that may predate alcohol initiation, emerge following initiation, or both (Spear, 2018). Work has identified neuroimaging correlates in adolescents before initiation that predict earlier AFAU, including reduced cortical thickness in frontal regions (Miller et al., 2024). Prospective studies have also identified differences in large-scale brain network recruitment before substance use initiation, including altered network engagement during spatial memory and emotional inhibitory control among adolescents who subsequently initiated substance use (Cohen-Gilbert et al., 2025; Sneider et al., 2025). Furthermore, prefrontal cortex (PFC) surface area has been shown to be reduced following initiation of alcohol use (Infante et al., 2018), supporting preclinical work that shows adolescent alcohol exposure itself can produce persistent neurobiological and behavioral alterations that remain evident in adulthood (Crews et al., 2019). The PFC undergoes prolonged development through adolescence into young adulthood, with frontal myelination and synaptic organization continuing into the third decade of life (Petanjek et al., 2011; Sowell et al., 2003). This prolonged developmental window raises the possibility that AFAU-related correlates may persist, evolve, or interact with ongoing brain development into young adulthood.

Although previous work has linked AFAU with adolescent brain characteristics, it remains unclear whether AFAU-related brain patterns persist into young adulthood and how such patterns relate to adult substance use behavior. Young adulthood is a particularly relevant period in which to examine these associations because frontal cortical structure and functional networks continue to mature alongside increasing independence and opportunities for substance use. Here, we leveraged the Human Connectome Project Young Adult (HCP-YA) cohort, which includes multimodal neuroimaging data from adults aged 22–35 years (Van Essen et al., 2013), to test whether brain signatures associated with AFAU are detectable in young adulthood and how these signatures relate to subsequent drinking behaviors.

While gray matter morphology has been extensively studied in relation to AFAU, the role of gray matter (intracortical) myelin remains relatively unexplored. Intracortical myelin (ICM) is an important component of cortical organization and has been implicated in healthy brain development as well as neurological and psychiatric disorders, in part through its contribution to neural signaling and cortical network organization (Sprooten et al., 2019). Importantly, ICM follows a graded pattern of cortical organization, with higher levels and earlier developmental changes in primary sensory and motor regions and more gradual maturation toward higher-order association regions (Baum et al., 2022). This sensorimotor-to-association gradient reflects the prolonged and regionally heterogeneous development of cortical microstructure across adolescence and into early adulthood, making ICM a potentially sensitive measure of developmental differences associated with AFAU. Despite its potential importance, ICM has received little attention in alcohol research (Rice & Gu, 2019), limiting our understanding of how AFAU and subsequent alcohol use may relate to cortical development beyond conventional measures of thickness and surface area.

In addition to structural alterations, AFAU has been associated with differences in functional brain organization. Earlier AFAU has been associated with greater functional connectivity (FC) between frontal regions (Nguyen-Louie et al., 2018), as well as alterations in large-scale brain networks, including the Default Mode and Salience Networks (Kardan et al., 2025). The Default Mode Network (DMN), anchored in medial PFC, posterior cingulate, precuneus and temporoparietal cortex, supports internally oriented processes including self-referential cognition, autobiographical memory, social cognition, future-oriented cognition, and valuation (Hiser & Koenigs, 2018; Menon, 2023). The Salience Network (SN), anchored in insula, anterior cingulate, and lateral PFC, contributes to the detection, integration, and prioritization of behaviorally relevant information, particularly emotionally and motivationally salient stimuli, and facilitates coordination between large-scale brain networks (Menon, 2023; Menon & Uddin, 2010). The DMN and SN together encompass prefrontal systems relevant to affective, reward, and salience processing, and are implicated in psychopathology (Menon, 2011), providing a framework for examining functional organization associated with earlier age of first alcohol use and subsequent alcohol drinking behaviors.

More broadly, evidence suggests that neurobiological correlates of AFAU are multimodal in nature, spanning cortical morphology, tissue microstructure, and brain network organization (Infante et al., 2018; Miller et al., 2024; Nguyen-Louie et al., 2018). However, most neuroimaging studies have examined these modalities independently, potentially overlooking coordinated patterns of variation across the brain. Because brain development reflects the interaction of multiple structural and functional processes, analytical approaches that integrate multimodal information may provide a more comprehensive understanding of the neural correlates of early alcohol exposure. Recent advances in multimodal imaging data fusion have enabled the identification of shared patterns of variation. SuperBigFLICA (SBF; Cheng et al., 2025; Gong et al., 2023) extends an earlier data-fusion approach, FMRIB’s Linked Independent Component Analysis (FLICA; Groves et al., 2011), by incorporating semi-supervised machine learning to identify multimodal patterns that are predictive of a target phenotype while retaining the ability to decompose large-scale imaging data into a latent space enriched with target-related information that may generalize to other related behaviors. This framework is therefore well suited to investigating multimodal neurobiological patterns associated with AFAU and their relationships to subsequent alcohol use behaviors.

The present study used data from HCP-YA to first identify multimodal brain covariance patterns using SBF that are maximally predictive of AFAU, integrating cortical thickness, surface area, ICM, and resting-state FC of frontal subnetworks of the DMN and SN. Next, we used the resulting multimodal latent space enriched for information related to AFAU to identify multimodal brain profiles associated with different patterns of subsequent alcohol use. To examine whether this latent space generalized to other substance use behaviors, we also examined how latent spatial patterns relate to age of first cannabis use and lifetime cannabis use.

## Methods

### Participants

Data were obtained from the open access Human Connectome Project Young Adult (HCP-YA; https://balsa.wustl.edu) study, a large community-based neuroimaging cohort of healthy adults aged 22–35 years (Van Essen et al. 2013). Participants with complete multimodal neuroimaging data and who reported age of first alcohol use (AFAU) information were included. HCP-YA enrollment excluded severe psychiatric, neurological, and medical conditions, resulting in a generally healthier sample with lower rates of substance use pathology than population-based cohorts. Detailed recruitment and exclusion procedures have been described in Van Essen et al. (2013).

### Clinical Measures

HCP-YA collected self-report substance use data via the Semi-Structured Assessment for the Genetics of Alcoholism (SSAGA; Bucholz et al., 1994). Self-report age of first alcohol use (AFAU) was the primary target variable for multimodal data fusion. Additional variables were examined in follow-up analyses to characterize adult alcohol and cannabis use behavior patterns, including drinking frequency during the previous 12 months (SSAGA_Alc_12_Frq), binge drinking frequency (≥5 drinks per occasion) during the previous 12 months (SSAGA_Alc_12_Frq_5plus), drinking frequency during the participant’s lifetime period of heaviest alcohol consumption (SSAGA_Alc_Hvy_Frq), binge drinking frequency during the lifetime period of heaviest alcohol consumption (SSAGA_Alc_Hvy_Frq_5plus), age of first cannabis use, and lifetime cannabis use (SSAGA_Mj_Times_Used). Non-binned alcohol and cannabis age of first use data were obtained directly from the HCP study team.

### Neuroimaging

MRI data were acquired on a Siemens 3T Skyra using standardized HCP-YA protocols (Glasser et al., 2013). Minimally pre-processed functional MRI data underwent group ICA using FSL’s MELODIC to generate subject-level network connectivity maps as described in https://humanconnectome.org/storage/app/media/documentation/s1200/HCP1200-DenseConnectome+PTN+Appendix-July2017.pdf. Structural MRI underwent FreeSurfer processing to generate cortical thickness, mid-thickness surface area, and ICM. Additional details in https://humanconnectome.org/storage/app/media/documentation/s1200/HCP_S1200_Release_Reference_Manual.pdf and Supplemental Methods.

#### Network selection

We focused our network selection on a parsimonious subset of networks spanning prefrontal cortical areas implicated in affective, salience and reward processing: the ventromedial frontal subnetwork of the DMN and the frontal subnetwork of SN (Supplemental Figure 1). These networks are anchored in frontal cortex and are highly relevant to AFAU (Kardan et al., 2025; Nguyen-Louie et al., 2018). These networks overlap with Yeo-17 Default C and Salience B respectively (Supplemental Figure 2; additional rationale in Supplemental Methods).

### Brain Imaging Data Fusion

The final imaging dataset consisted of three structural modalities, maps of cortical thickness (CT), mid-thickness surface area (SA), and intracortical myelin (ICM), and two functional connectivity (FC) modalities, the FC maps of the frontal subnetworks of the DMN and SN for each participant. These data were integrated using SuperBigFLICA (SBF; Cheng et al., 2026; Gong et al., 2023) to identify latent spatial patterns of shared variation across the modalities and corresponding subject loadings that are maximally predictive of AFAU. SBF was implemented using our open-access Python-based tool (Cheng et al., 2026; https://github.com/ANSR□laboratory/SuperBigFLICA_McL).

Training, testing, and external validation set creation details can be found in Supplemental Methods. The train and test datasets were used to identify optimal hyperparameters, including SBF component dimensionality (or number of components estimated; d=25 vs d=50), batch size (24-60, intervals of 12), and training epochs (10-50, intervals of 10).

AFAU served as the target variable with hyperparameter selection based on maximizing the correlation between predicted and observed AFAU in the test dataset. Following optimization, the selected final model was applied to the independent external validation dataset to assess generalizability.

**Model interpretability** was assessed using SHapley Additive exPlanations (SHAP; Lundberg & Lee, 2017; https://github.com/shap/shap) to quantify the contribution of individual SBF components to model predictions. Components were ranked by their mean absolute SHAP values, and the five most important multimodal components were selected for subsequent analyses.

#### Spatial pattern interpretability

Dice coefficients and percent overlap with the Yeo-17 parcellation were calculated separately for the positive and negative spatial maps of the top five SBF components (Yeo et al., 2011) using the Ciftify Python package (https://github.com/edickie/ciftify). Overlap with the Yeo networks was used only to aid interpretation.

### Brain-Behavior Profiles

To investigate associations between AFAU-related brain patterns and adult alcohol and cannabis use behaviors, canonical correlation analysis (CCA; Wang et al., 2020) was performed using Python’s Scikit-learn. CCA was used to identify maximally correlated brain-behavior latent profiles. The neuroimaging feature set consisted of participant loadings from the five highest-ranking SBF components identified by SHAP. The behavioral feature set included AFAU, cannabis first-use age, drinking frequency during the previous 12 months, binge drinking frequency during the previous 12 months, drinking frequency during the lifetime heaviest drinking period, binge drinking frequency during the lifetime heaviest drinking period, and lifetime cannabis use. Scikit-learn’s CCA function scales both the brain and behavior features before performing the analysis. Age, sex, and education were not included in the CCA, but their associations were assessed using post hoc correlations with the CCA modes (Smith et al., 2015).

**Statistical significance** of each canonical mode was evaluated via permutation testing using 1,000 permutations. Only modes that survived permutation testing were retained for interpretation. Canonical loadings and variable contributions were examined to characterize multimodal brain–behavior profiles associated with substance use patterns in adulthood, with canonical loadings interpreted as the strength of association between each observed variable and its corresponding latent dimension.

## Results

### Participants

622 participants (52.1% female, age=28.40±3.83 years, education=15.00±1.73 years) had complete imaging and clinical data. See Figure 1A for AFAU and Figure 1B-D for alcohol and cannabis use behaviors in the overall sample. See Table 1 for sample characteristics for the training, testing, and external validation splits. The splits showed no significant differences in age (F(2, 619) = 0.23, p = 0.79) or sex (χ^2^(2) = 0.30, p = 0.86).

**Figure 1.**
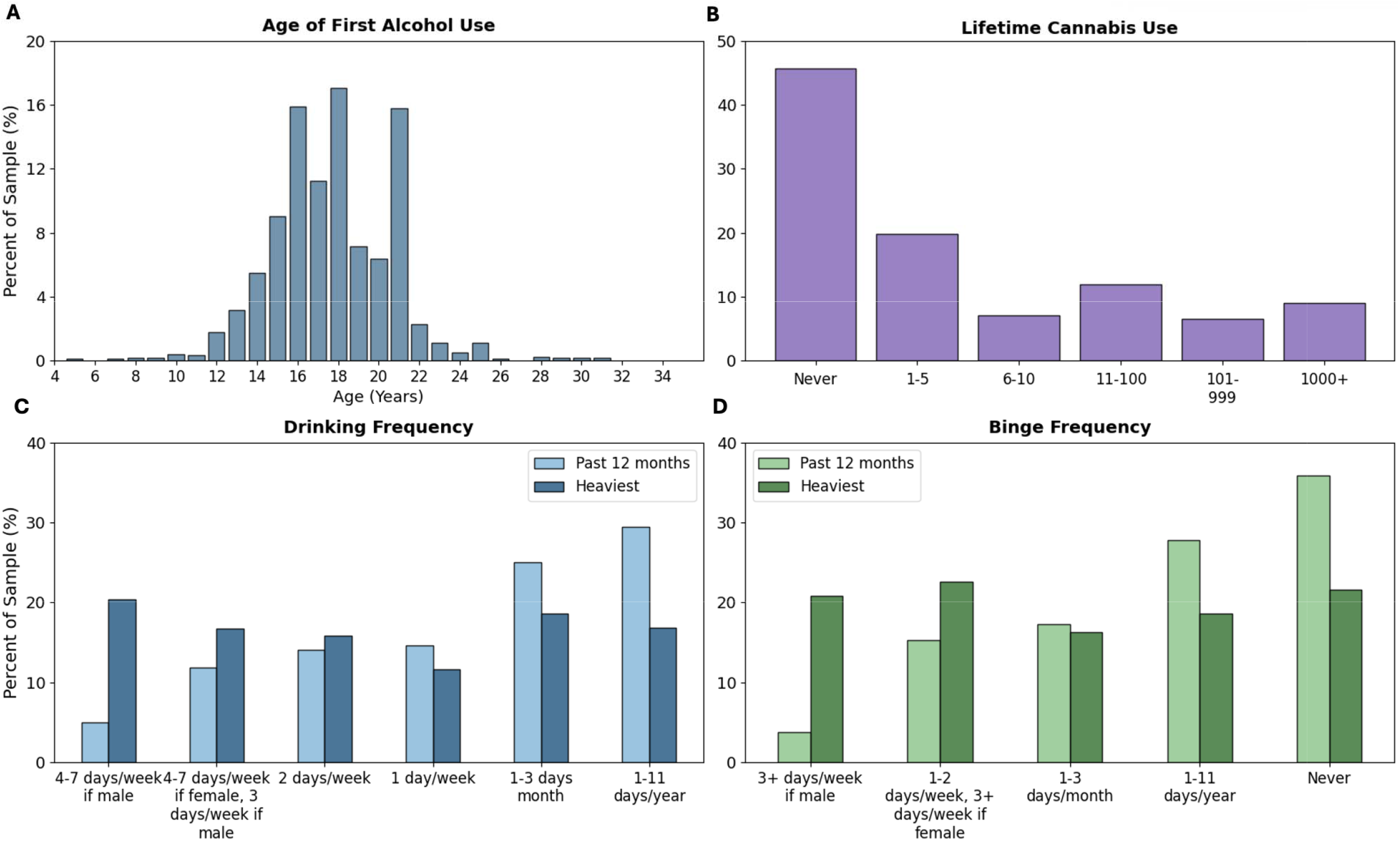
Overall sample substance-use profile. Percent of sample shown for **(A)** age of first alcohol use, **(B)** lifetime cannabis use (total times used), **(C)** drinking frequency (past 12 months vs. heaviest use), and **(D)** binge frequency (past 12 months vs. heaviest use).

**Table 1.**
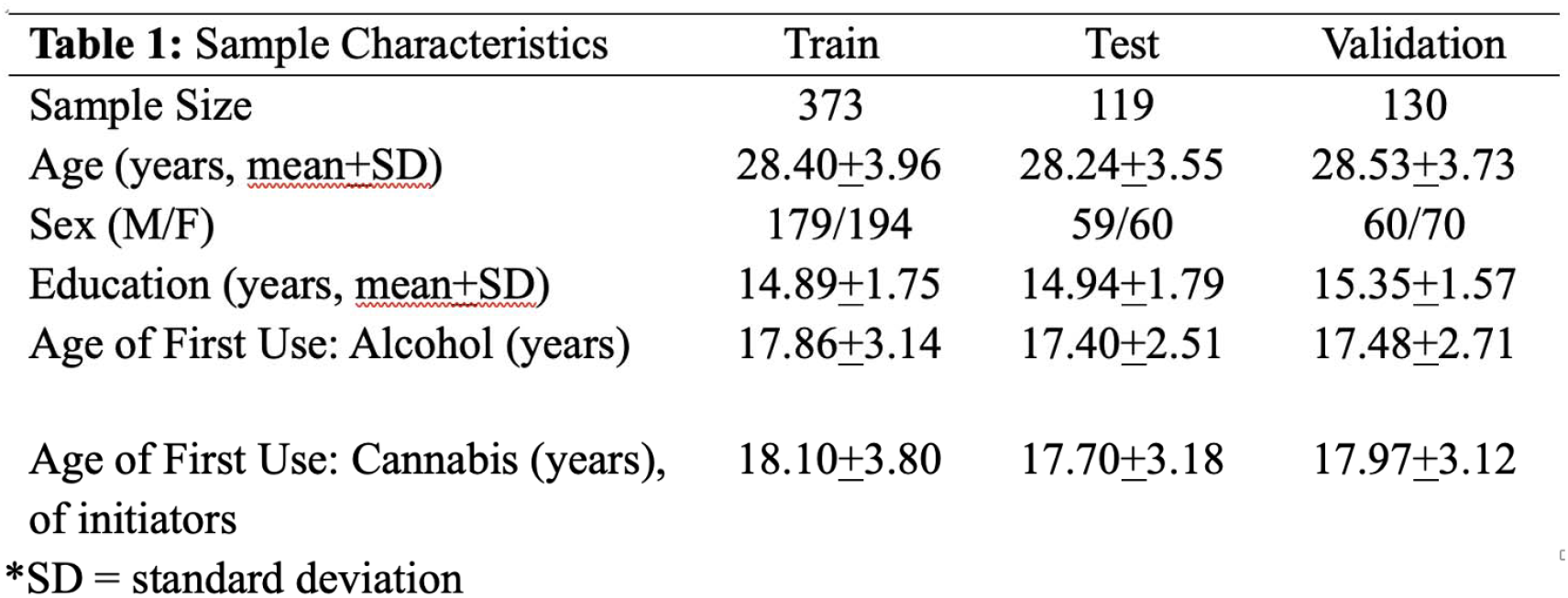
Demographics of the train, test, external validation splits. Cannabis age of first use is calculated from those who initiated.

| **Table 1: Sample Characteristics** | **Train** | **Test** | **Validation** |
| --- | --- | --- | --- |
| Sample Size | 373 | 119 | 130 |
| Age (years, <u>mean+SD</u>) | 28.40±3.96 | 28.24±3.55 | 28.53±3.73 |
| Sex (M/F) | 179/194 | 59/60 | 60/70 |
| Education (years, <u>mean+SD</u>) | 14.89±1.75 | 14.94±1.79 | 15.35±1.57 |
| Age of First Use: Alcohol (years) | 17.86±3.14 | 17.40±2.51 | 17.48±2.71 |
| Age of First Use: Cannabis (years),<br>of initiators | 18.10±3.80 | 17.70±3.18 | 17.97±3.12 |

\*SD = standard deviation

### Multi-modal Brain Patterns Maximally Predictive of Age of First Alcohol Use

Model optimization identified an optimal dimensionality of *d*=50 with a batch size of 48, reaching convergence within 15 training epochs. This solution demonstrated modest predictive performance (correlation between true and predicted initiation age) in both the held-out test sample (*r*=0.18) and the external validation (*r*=0.22), supporting the reproducibility and generalizability of the identified multimodal patterns sample SHAP analysis identified components 11, 25, 35, 18, and 45 as the strongest contributors to AFAU prediction (Figure 2A). Examination of modality contributions revealed both unimodal and multimodal components, with components 11 and 45 being highly multimodal (Figure 2B). Per-subject SHAP scores color-coded by subject loading can be found in Figure 2C, capturing the direction of effect for each SBF component. SBF loading versus SHAP score scatter plots can be found in Supplemental Figure 3, showing that greater expression of Components 11, 18, and 45 predicts later AFAU, while greater expression of Components 25 and 35 predicts earlier AFAU.

**Figure 2.**
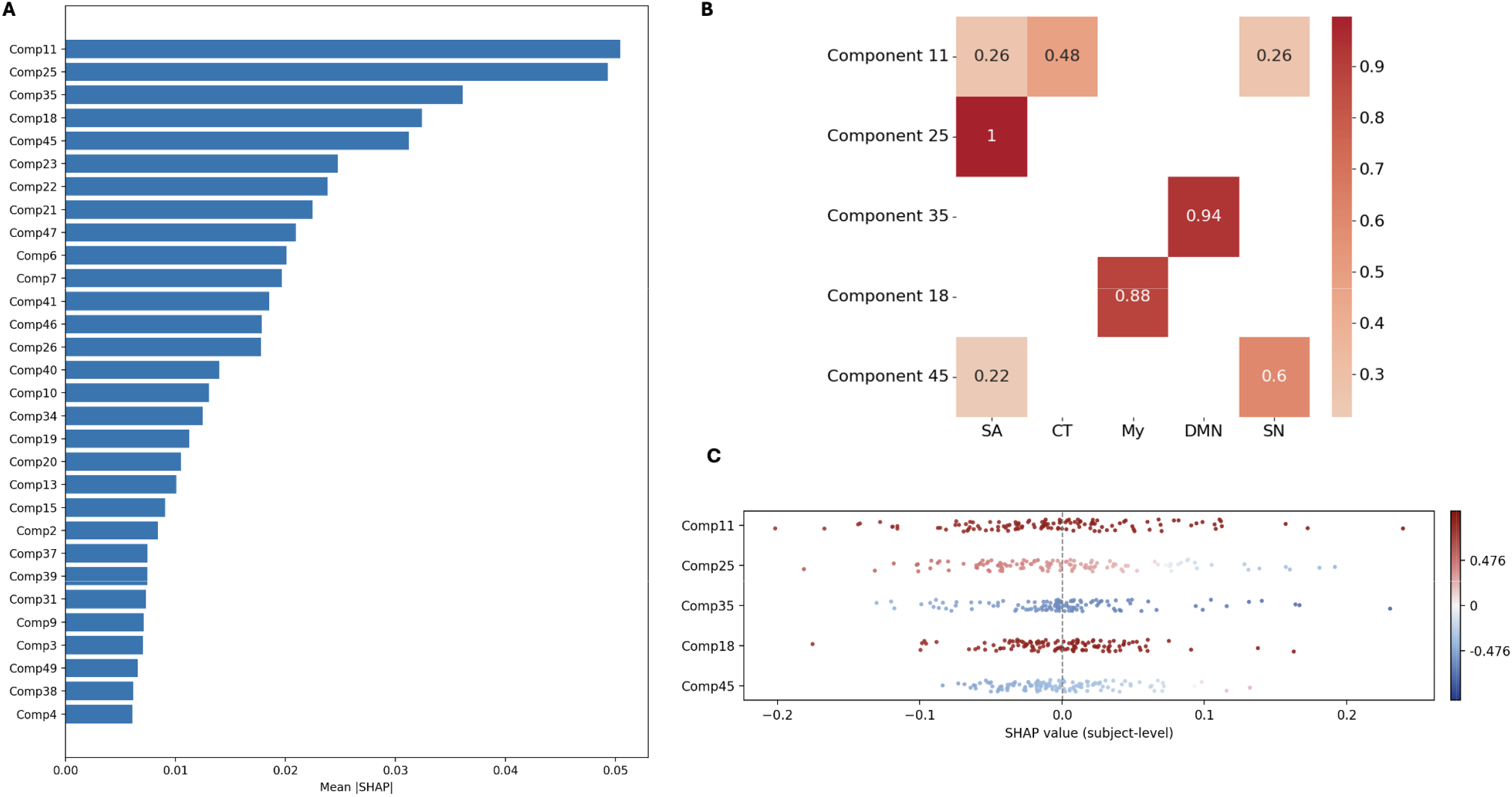
**A**. Mean |SHAP| score for each SBF component, ordered from largest to smallest |SHAP|. **B**. Heatmap of modality weights showing contributions to the top 5 SBF components, thresholded at 0.2. **C**. SHAP value for each participant for the top 5 SBF components, colored by SBF subject loading.

The spatial maps of the top 5 components can be found in Figure 3, thresholded to show the top 10% of spatial map values that reflect the strongest signals (top 5% each of negative and positive tails). To aid interpretation of SBF spatial maps, we overlaid resulting thresholded maps onto the Glasser MSMAll fine parcellation (Glasser et al., 2013; Supplemental Figure 4) and computed spatial overlap using Dice coefficients and percent overlap with the Yeo-17 network parcellation (Yeo et al., 2011); results are shown in Figure 4.

**Figure 3.**
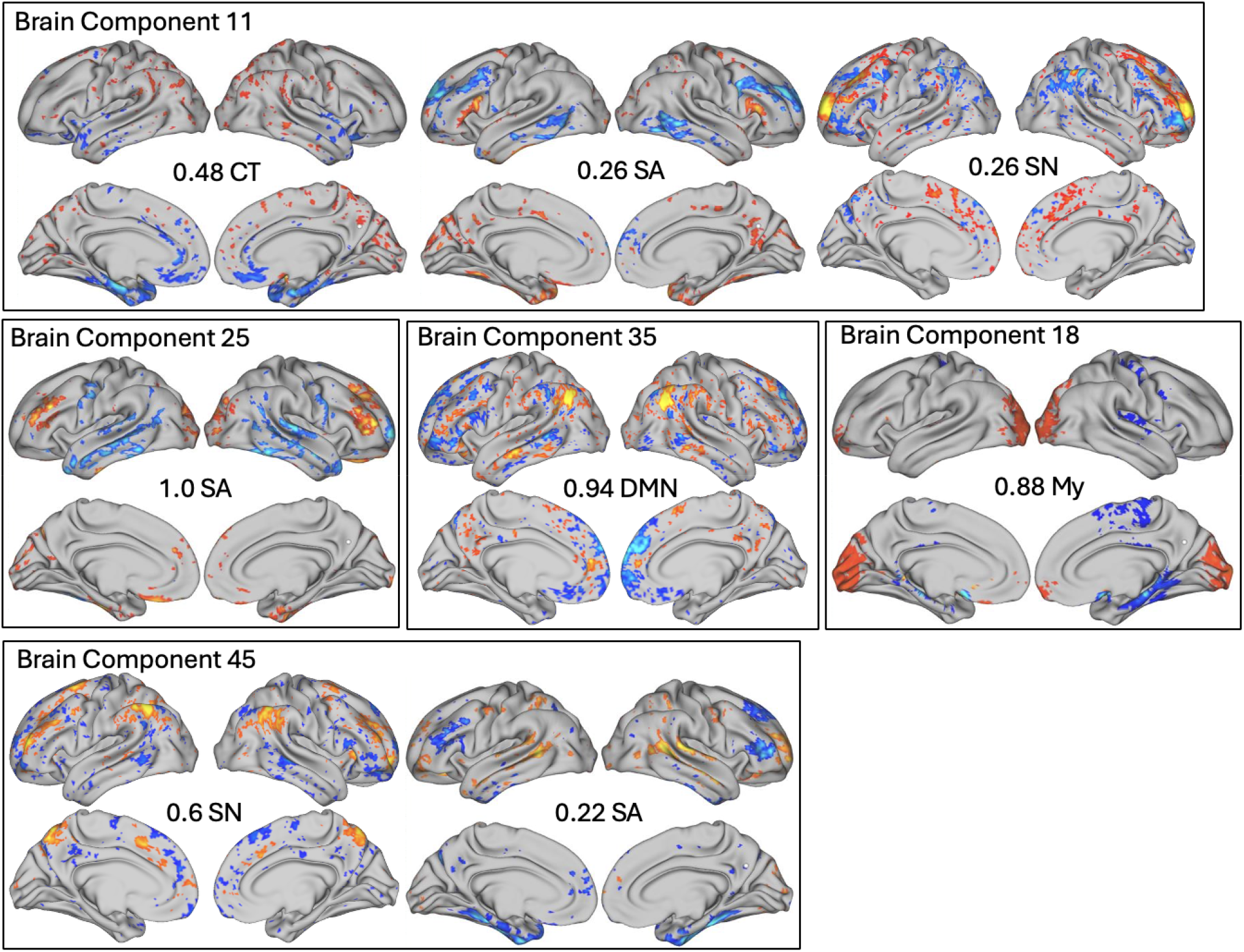
Spatial maps of the top 5 SBF components as ranked by mean SHAP score. Components 11 and 45 show multimodal linked covariance patterns, while components 25, 35 and 18 are unimodal. SA = Surface Area, SN = frontal subnetwork of the Salience Network, CT = cortical thickness, DMN = Default Mode Network, My = intracortical myelin.

Component 11, the most important feature for predicting AFAU, with higher loadings predicting later AFAU, reflected coordinated variation across cortical thickness, surface area, and resting state FC of SN. Lower CT in insula and medial and inferior temporal cortex but higher SA in the same temporal areas, was linked to FC of SN within the network and with areas in dorsolateral PFC (dlPFC) and posterior parietal cortex. As shown in Figure 4, these regions largely overlap with limbic and cognitive control networks of the Yeo-17 parcellation. Altogether, this component represents a joint brain structure-function covariance pattern that links structural covariance (SC) in CT of areas overlapping with limbic and DMN networks and SA of areas overlapping limbic and control networks with SN FC within network and with areas implicated in cognitive control.

**Figure 4.**
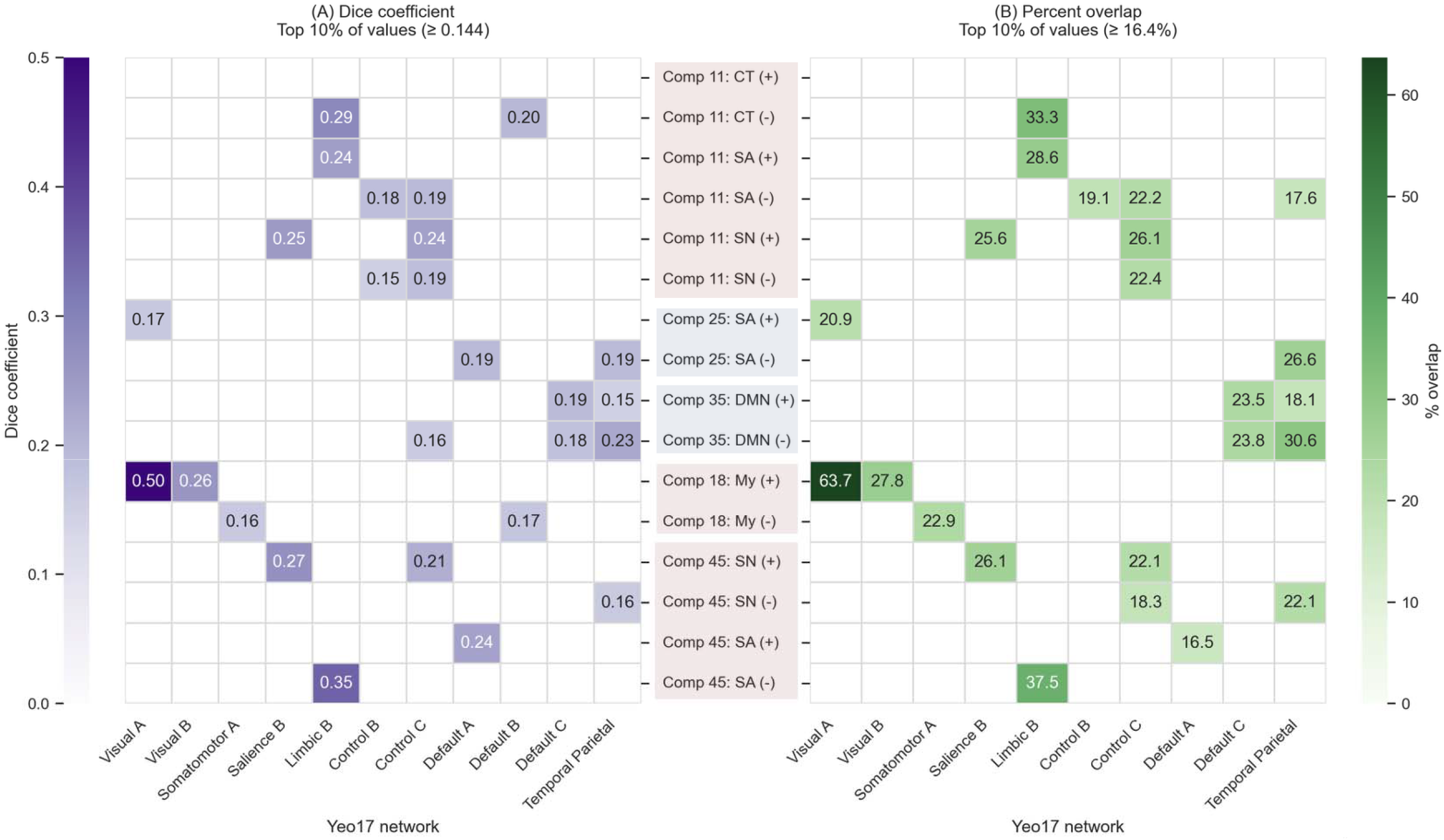
Dice coefficient in purple **(A)** and percent overlap in green **(B)** of the top 5 spatial maps with Yeo-17 atlas. Constraining to the top 10th percentile creates a threshold of 0.144 and 16.4%, respectively. Red shading of component labels (middle column) indicates that higher loadings on the component predicted later age of first use while higher loadings on blue shaded components predicted earlier AFAU.

The second component predictive of later AFAU is also a multimodal pattern, Component 45 -characterized by increased within-network FC of the SN linked to a structural covariance pattern of reduced SA of inferior temporal and anterior inferior frontal cortices and increased SA of middle and superior temporal gyri and orbitofrontal cortex (OFC), areas overlapping with DMN/limbic networks. Notably, the DMN FC modality also contributed weakly to this component (modality weight = 0.10), showing alterations of DMN with areas in SN (visualized in Supplemental Figure 4). Altogether, this component reflects a joint SC-FC pattern linking SN FC with structural and functional covariance of emotion/reward areas and areas implicated in attention/salience.

Component 18, also associated with later AFAU, reflected a structural covariance pattern of ICM consisting of increased ICM in the visual cortex and decreased myelin in somatomotor cortex.

Two components in which higher loadings predicted earlier AFAU were also identified. Component 25 is a unimodal component reflecting lower SA of middle temporal gyrus and superior medial PFC (regions overlapping Yeo-17’s temporoparietal network) covarying with greater SA of visual cortex, OFC, and dlPFC. Therefore, this component represents temporoparietal/visual SA alterations that predict earlier AFAU. Component 35 is a unimodal pattern of DMN FC with inferior parietal cortex, medial PFC, and middle temporal gyrus. This component largely overlaps with Yeo-17 temporoparietal and Default C networks. Together, this component represents FC between DMN sub-networks and temporoparietal areas implicated in higher-order processing and integration of information.

### Differential Brain Patterns Associated with Substance Use Profiles

To characterize relationships between AFAU-related brain patterns and adult alcohol and cannabis use behaviors, CCA was used to link loadings on the five most important data fusion patterns to key alcohol and cannabis use variables in a multivariate analysis. We identified three significant multimodal CCA brain– behavior modes based on permutation testing (Mode 1: *r* = 0.2750, *p* = 0.014; Mode 2: *r* = 0.2249, *p* = 0.005; Mode 3: *r* = 0.1568, *p* = 0.029; Supplemental Figure 5). Variable loadings on the brain and behavioral canonical variates (or modes) indicating the strongest contributors to each mode are shown in Figure 5 (unthresholded loadings are shown in Supplemental Figure 6). We evaluated age, sex, and education as covariates using post hoc correlations with subject loadings on each CCA mode. Correlations indicated weak associations of these covariates with all modes (mode 1/2/3: age r=0.08/-0.14/-0.07, sex r=-0.16/-0.04/-0.06, education r=-0.08/-0.13/0.17).

**Figure 5.**
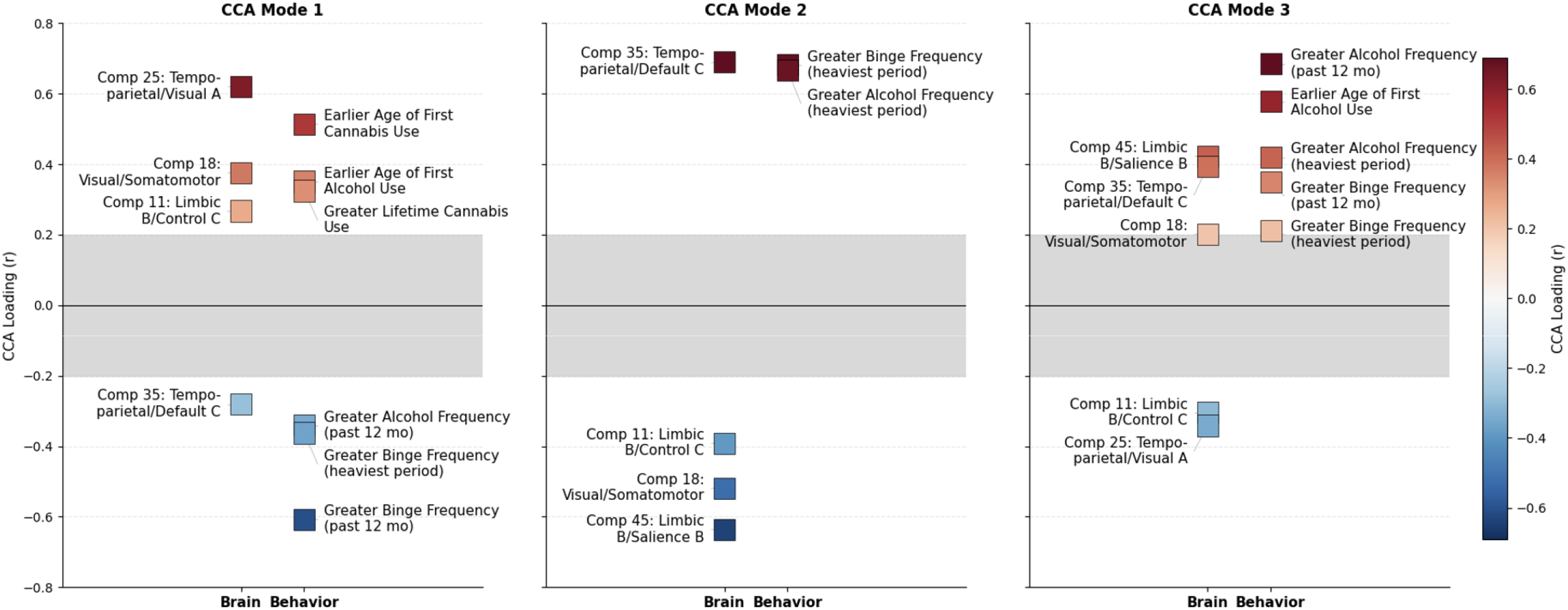
CCA loadings for the 3 significant modes, thresholded at 0.2. Signs of loadings in Mode 2 were flipped for easier interpretation. dlPFC-OFC-Temp-Vis SA = dorsolateral prefrontal cortex-temporal-visual Surface Area; GM My = intracortical myelin; DMN/SN FC = Default Mode Network/Salience Network Functional Connectivity.

Mode 1 reflects an axis contrasting cannabis and alcohol behaviors. Although alcohol and cannabis use are often positively correlated at the population level, the first canonical mode identified a latent dimension that differentiated individuals with earlier age of first use of cannabis and alcohol and greater lifetime cannabis use on one end and greater recent and past binge drinking and recent frequent drinking on the other end. This behavioral variate was linked to a brain profile of greater representation of components 25, 18, and 11 at one end and lower expression of component 35 at the other end. Together, these profiles suggest a timing- and substance-specific pattern in which earlier initiation and greater lifetime cannabis use, with lower binge drinking, were associated with greater representation of 25, 18 and 11 and lower expression of component 35. Conversely, later initiation and greater binge drinking were associated with greater expression of component 35 and lower expression of components 25, 18 and 11. Components 25 and 18, the strongest positive contributors to this mode, both reflect structural covariance of the visual cortex through greater SA and ICM density, respectively. Component 25 also captures reduced SA in regions overlapping the temporoparietal network, whereas the strongest negative contributor, Component 35, reflects FC of the DMN, both within-network and with temporoparietal areas. Thus, Mode 1 contrasts an early initiation, higher-lifetime-cannabis-use profile associated with greater visual cortex SC and lower DMN FC with a later initiation, greater-binge-drinking profile associated with greater DMN FC and lower visual cortical SC.

Mode 2 reflects a brain profile linked to alcohol drinking during the individual’s heaviest drinking period that was not related to age of first use of either alcohol or cannabis. Greater drinking frequency and binge drinking during the heaviest drinking period were associated with increased expression of component 35 (DMN FC) but reduced expression of components 45 (SN FC linked to limbic SA), 11 (limbic and cognitive control CT/SN FC), and 18 (primary cortex ICM). Thus, this mode represents a historical drinking profile associated with DMN FC, limbic CT, and ICM in primary cortex.

Mode 3 reflects a brain-behavior mode of greater alcohol involvement, including earlier AFAU, more frequent drinking, and greater binge drinking (past year and during the heaviest drinking period), associated with greater expression of components 45, 35, and 18 and lower expression of components 25 and 11. Conversely, individuals with later age of first alcohol use and lower levels of alcohol use had stronger expression of components 25 and 11 and lower expression of components 45, 35, and 18. Mode 3 links heavier alcohol use and binge drinking to a system-level pattern characterized by greater expression of SN FC linked to limbic SA (Component 45) and FC within the DMN (Component 35), coupled with reduced expression of SN FC linked to limbic and cognitive control morphometry (Component 11) and dlPFC-temporal-visual SA (Component 25).

## Discussion

In this study, we applied a semi-supervised multimodal imaging data fusion approach to investigate cortical morphometry and prefrontal FC linked to age of first use of alcohol. This resulted in latent multimodal brain patterns predictive of AFAU, and linked to alcohol and cannabis use more generally. Importantly, the CCA modes suggested partially dissociable neurobiological patterns associated with different profiles of alcohol and cannabis use behaviors.

We found that variation in AFAU across the sample is connected to coordinated variation across structural and functional systems, including thickness and surface area of cortex, ICM, and PFC-anchored network-level organization. Two unimodal components were identified that were predictive of earlier AFAU. Component 25, the largest predictor of earlier AFAU, is a pattern of dlPFC, OFC, and temporal cortex SA alterations, while Component 35 is a pattern of anti-covariance between frontal and posterior parietal areas of the DMN. These findings align with the large body of literature that identifies associations between earlier AFAU and structural reductions in frontal cortex, particularly dlPFC and OFC (Infante et al., 2018; Miller et al., 2024; Spear, 2018), and DMN alterations (Kardan et al., 2025; Nguyen-Louie et al., 2018).

Prospective adolescent studies have also identified differences in large-scale brain network recruitment prior to substance use initiation, including during spatial memory and emotional inhibitory control, suggesting that some network-level differences associated with later substance use may be present before initiation (Cohen-Gilbert et al., 2025; Sneider et al., 2025). Together, these findings raise the possibility that some of the AFAU-related patterns observed in young adulthood may reflect latent neurodevelopmental vulnerability present before substance use initiation, although the present design cannot distinguish such vulnerability from consequences of subsequent substance exposure.

The remaining top three components predict later AFAU. Component 11 was the greatest predictor of later AFAU and identified CT reductions in areas overlapping the limbic network, particularly the medial temporal lobe. Component 45 also identified structural medial temporal lobe alterations associated with later AFAU. These findings align with previous work (Miller et al., 2024) which found increased medial temporal lobe cortical thickness was associated with earlier AFAU. ICM of the primary cortex emerged as an important contributing brain feature (Component 18), extending prior work that has largely focused on more traditional cortical morphometric measures and suggesting that ICM-related microstructural variation also reflects AFAU-related brain differences.

The canonical correlation analysis showed that these multimodal brain signatures map onto distinct dimensions of substance use behavior rather than a single generalized risk factor. The first mode captured a contrast between cannabis- and alcohol-related behaviors, despite their typical positive association at the population level. This axis reflected a continuum in which greater cannabis burden and lower alcohol burden were associated with alterations in primary cortices, whereas greater alcohol burden and lower cannabis burden were associated with differences in DMN connectivity. The second mode linked heavier alcohol consumption during peak drinking periods to DMN organization, characterized by reduced FC of frontal versus increased FC of posterior DMN nodes. This pattern suggests that cumulative alcohol exposure may be preferentially associated with DMN connectivity independent of when alcohol was first used. The third mode reflected a greater alcohol involvement profile that juxtaposed SN connectivity differences with structural limbic changes. In all, we identified distinct multivariate brain patterns associated with different alcohol and cannabis behavior profiles later in life.

There are some limitations of our study. First, the cross-sectional and retrospective design precludes inference about whether these brain patterns precede or follow substance use initiation. Longitudinal imaging beginning before initiation will be essential to disentangle latent vulnerability from consequences of subsequent substance exposure. Additionally, although HCP-YA provides high-quality multimodal imaging, its relatively limited substance use phenotyping constrains finer-grained interpretation of substance use behavior. Future work leveraging datasets like the Adolescent Brain Cognitive Development cohort, which includes longitudinal imaging before and after substance use initiation and in-depth behavioral phenotyping, is needed to differentiate the neurodevelopmental correlates of early initiation from those associated with continued heavy use versus limited subsequent use.

In conclusion, this study identified multimodal brain covariance patterns associated with age of first use of alcohol that are observable in young adulthood. Our findings provide support for prior research showing associations between age of first use of alcohol and prefrontal FC and structural differences. Our approach integrates cortical morphometry and ICM with DMN and SN network FC, highlighting linked covariance among these measures across structural and functional systems. We also identified ICM as a novel contributor to alcohol first-use related brain variation in young adults. Furthermore, distinct profiles of these multimodal brain patterns were associated with different profiles of alcohol and cannabis behaviors, distinguishing alcohol use-related brain-behavior patterns from those related to cannabis use. Together, these findings highlight the value of a multivariate, multimodal perspective for revealing distinct neurobiological profiles associated with the timing and patterns of substance use.

## Data Availability

All data produced in the present study are available upon reasonable request to the authors.

## Notes

### Competing Interest Statement

The authors have declared no competing interest.

### Author Declarations

Human Connectome Project Young Adult: https://balsa.wustl.edu/

### Funding

- National Institute on Drug Abuse: 5T32DA015036-24

## References

Baum, G. L., Flournoy, J. C., Glasser, M. F., Harms, M. P., Mair, P., Sanders, A. F. P., Barch, D. M., Buckner, R. L., Bookheimer, S., Dapretto, M., Smith, S., Thomas, K. M., Yacoub, E., Van Essen, D. C., & Somerville, L. H. (2022). Graded Variation in T1w/T2w Ratio during Adolescence: Measurement, Caveats, and Implications for Development of Cortical Myelin. The Journal of Neuroscience: The Official Journal of the Society for Neuroscience, 42(29), 5681–5694. 10.1523/JNEUROSCI.2380-21.2022

Ben Shalom, D., & Skandalakis, G. P. (2025). Four Streams Within the Prefrontal Cortex: Integrating Structural and Functional Connectivity. The Neuroscientist: A Review Journal Bringing Neurobiology, Neurology and Psychiatry, 31(1), 8–13. 10.1177/10738584241245304

Brennan, M. M., Mongan, D., Doyle, A., Millar, S. R., Cavallaro, M., Zgaga, L., Smyth, B. P., Nixon, E., Ivers, J.-H., Galvin, B., Walsh, C., McCrory, C., & McCarthy, N. D. (2025). Early and risky adolescent alcohol use independently predict alcohol, tobacco, cannabis and other drug use in early adulthood in Ireland: A longitudinal analysis of a nationally representative cohort. BMC Public Health, 25(1), 1129. 10.1186/s12889-025-22262-w

Bucholz, K. K., Cadoret, R., Cloninger, C. R., Dinwiddie, S. H., Hesselbrock, V. M., Nurnberger, J. I., Reich, T., Schmidt, I., & Schuckit, M. A. (1994). A new, semi-structured psychiatric interview for use in genetic linkage studies: A report on the reliability of the SSAGA. Journal of Studies on Alcohol, 55(2), 149–158. 10.15288/jsa.1994.55.149

Cheng, Y., Medina, A., Korponay, C., Beckmann, C. F., Harper, D., Nickerson, L., & Alzheimer’s Disease Neuroimaging Initiative. (2026). Investigating the amyloid-tau-neurodegeneration framework in Alzheimer’s disease using semi-supervised multimodal imaging data fusion. Alzheimer’s & Dementia, 18(2), e70360. 10.1002/dad2.70360

Cheng, Y., Medina, A., Korponay, C., Beckmann, C. F., Harper, D., Nickerson, L., & the Alzheimer’s Disease Neuroimaging Initiative. (2025). Investigating the Amyloid-Tau-Neurodegeneration Framework in Alzheimer’s Disease Using Semi-Supervised Multimodal Imaging Data Fusion. medRxiv, 2025.12.11.25341830. 10.64898/2025.12.11.25341830

Cohen-Gilbert, J. E., Sneider, J. T., Oot, E. N., Seraikas, A. M., Schuttenberg, E. M., Harris, S. K., Nickerson, L. D., & Silveri, M. M. (2025). Risk for Adolescent Substance Use Initiation: Associations with Large-Scale Brain Network Recruitment During Emotional Inhibitory Control. Behavioral Sciences, 15(10), 1407. 10.3390/bs15101407

Crews, F. T., Robinson, D. L., Chandler, L. J., Ehlers, C. L., Mulholland, P. J., Pandey, S. C., Rodd, Z. A., Spear, L. P., Swartzwelder, H. S., & Vetreno, R. P. (2019). Mechanisms of Persistent Neurobiological Changes Following Adolescent Alcohol Exposure: NADIA Consortium Findings. Alcoholism, Clinical and Experimental Research, 43(9), 1806–1822. 10.1111/acer.14154

Flores-López, M., Reviriego, R., García-Marchena, N., Requena-Ocaña, N., Ruiz-Ruiz, J. J., Herrera-Imbroda, J., Bordallo, A., Suarez, J., Moreno-Peral, P., Bellón, J. Á., de Fonseca, F. R., Pavón-Morón, F. J., & Serrano, A. (2026). Developmental Timing of Alcohol Initiation as a Marker of Adult Clinical Complexity in Alcohol Use Disorder: A Cross-Sectional Study. Medical Sciences, 14(5), 535. 10.3390/medsci14050535

Gardner, L. A., Stockings, E., Champion, K. E., Mather, M., & Newton, N. C. (2024). Alcohol initiation before age 15 predicts earlier hazardous drinking: A survival analysis of a 7-year prospective longitudinal cohort of Australian adolescents. Addiction, 119(3), 518–529. 10.1111/add.16376

Glasser, M. F., Sotiropoulos, S. N., Wilson, J. A., Coalson, T. S., Fischl, B., Andersson, J. L., Xu, J., Jbabdi, S., Webster, M., Polimeni, J. R., Van Essen, D. C., & Jenkinson, M. (2013). The minimal preprocessing pipelines for the Human Connectome Project. NeuroImage, 80, 105–124. 10.1016/j.neuroimage.2013.04.127

Glasser, M. F., & Van Essen, D. C. (2011). Mapping human cortical areas in vivo based on myelin content as revealed by T1- and T2-weighted MRI. The Journal of Neuroscience: The Official Journal of the Society for Neuroscience, 31(32), 11597–11616. 10.1523/JNEUROSCI.2180-11.2011

Gong, W., Bai, S., Zheng, Y.-Q., Smith, S. M., & Beckmann, C. F. (2023). Supervised Phenotype Discovery From Multimodal Brain Imaging. IEEE Transactions on Medical Imaging, 42(3), 834–849. 10.1109/TMI.2022.3218720

Groves, A. R., Beckmann, C. F., Smith, S. M., & Woolrich, M. W. (2011). Linked independent component analysis for multimodal data fusion. NeuroImage, 54(3), 2198–2217. 10.1016/j.neuroimage.2010.09.073

Harms, M. P., Somerville, L. H., Ances, B. M., Andersson, J., Barch, D. M., Bastiani, M., Bookheimer, S. Y., Brown, T. B., Buckner, R. L., Burgess, G. C., Coalson, T. S., Chappell, M. A., Dapretto, M., Douaud, G., Fischl, B., Glasser, M. F., Greve, D. N., Hodge, C., Jamison, K. W., … Yacoub, E. (2018). Extending the Human Connectome Project across ages: Imaging protocols for the Lifespan Development and Aging projects. NeuroImage, 183, 972–984. 10.1016/j.neuroimage.2018.09.060

Hiser, J., & Koenigs, M. (2018). The multifaceted role of ventromedial prefrontal cortex in emotion, decision-making, social cognition, and psychopathology. Biological Psychiatry, 83(8), 638–647. 10.1016/j.biopsych.2017.10.030

Infante, M. A., Courtney, K. E., Castro, N., Squeglia, L. M., & Jacobus, J. (2018). Adolescent Brain Surface Area Pre- and Post-Cannabis and Alcohol Initiation. Journal of Studies on Alcohol and Drugs, 79(6), 835–843. 10.15288/jsad.2018.79.835

Johnston, L. D., O’Malley, P. M., Bachman, J. G., & Schulenberg, J. E. (2022). Monitoring the future: National survey results on drug use, 1975-2022: Secondary school students: (567272009-001) [Dataset]. 10.1037/e567272009-001

Kardan, O., Weigard, A. S., Cope, L. M., Martz, M. E., Angstadt, M., McCurry, K. L., Michael, C., Hardee, J. E., Hyde, L. W., Sripada, C., & Heitzeg, M. M. (2025). Functional Brain Connectivity Predictors of Prospective Substance Use Initiation and Their Environmental Correlates. Biological Psychiatry. Cognitive Neuroscience and Neuroimaging, 10(2), 203–212. 10.1016/j.bpsc.2024.10.002

Lundberg, S., & Lee, S.-I. (2017). A Unified Approach to Interpreting Model Predictions (Version 2). arXiv.10.48550/ARXIV.1705.07874

Menon, V. (2023). 20 years of the default mode network: A review and synthesis. Neuron, 111(16), 2469–2487. 10.1016/j.neuron.2023.04.023

Menon, V., & Uddin, L. Q. (2010). Saliency, switching, attention and control: A network model of insula function. Brain Structure and Function, 214(5–6), 655–667. 10.1007/s00429-010-0262-0

Miller, A. P., Baranger, D. A. A., Paul, S. E., Garavan, H., Mackey, S., Tapert, S. F., LeBlanc, K. H., Agrawal, A., & Bogdan, R. (2024). Neuroanatomical Variability and Substance Use Initiation in Late Childhood and Early Adolescence. JAMA Network Open, 7(12), e2452027. 10.1001/jamanetworkopen.2024.52027

Nguyen-Louie, T. T., Simmons, A. N., Squeglia, L. M., Infante, M. A., Schacht, J. P., & Tapert, S. F. (2018). Earlier Alcohol Use Onset Prospectively Predicts Changes in Functional Connectivity. Psychopharmacology, 235(4), 1041–1054. 10.1007/s00213-017-4821-4

Nickerson, L. D., Smith, S. M., Öngür, D., & Beckmann, C. F. (2017). Using Dual Regression to Investigate Network Shape and Amplitude in Functional Connectivity Analyses. Frontiers in Neuroscience, 11, 115. 10.3389/fnins.2017.00115

Petanjek, Z., Judaš, M., Šimić, G., Rašin, M. R., Uylings, H. B. M., Rakic, P., & Kostović, I. (2011).Extraordinary neoteny of synaptic spines in the human prefrontal cortex. Proceedings of the National Academy of Sciences, 108(32), 13281–13286. 10.1073/pnas.1105108108

Rice, J., & Gu, C. (2019). Function and Mechanism of Myelin Regulation in Alcohol Abuse and Alcoholism.BioEssays, 41(7), 1800255. 10.1002/bies.201800255

Smith, S., Nichols, T., Vidaurre, D., Winkler, A., Behrens, T., Glasser, M., Ugurbil, K., Barch, D., Van Essen, D., & Miller, K. (2015). A positive-negative mode of population covariation links brain connectivity, demographics and behavior. Nature Neuroscience, 18(11), 1565–1567. 10.1038/nn.4125

Sneider, J. T., Cohen-Gilbert, J. E., Oot, E. N., Seraikas, A. M., Schuttenberg, E. M., Stallman, A., Hamilton, D. A., Harris, S. K., Sabolek, H., Kumar, P., Nickerson, L. D., & Silveri, M. M. (2025). Brain network signatures of spatial memory in adolescents at risk for substance use. Alcohol, Clinical & Experimental Research, 49(10), 2172–2183. 10.1111/acer.70155

Sowell, E. R., Peterson, B. S., Thompson, P. M., Welcome, S. E., Henkenius, A. L., & Toga, A. W. (2003). Mapping cortical change across the human life span. Nature Neuroscience, 6(3), 309–315. 10.1038/nn1008

Spear, L. P. (2018). Effects of adolescent alcohol consumption on the brain and behaviour. Nature Reviews Neuroscience, 19(4), 197–214. 10.1038/nrn.2018.10

Sprooten, E., O’Halloran, R., Dinse, J., Lee, W. H., Moser, D. A., Doucet, G. E., Goodman, M., Krinsky, H., Paulino, A., Rasgon, A., Leibu, E., Balchandani, P., Inglese, M., & Frangou, S. (2019). Depth-dependent intracortical myelin organization in the living human brain determined by in vivo ultra-high field magnetic resonance imaging. NeuroImage, 185, 27–34. 10.1016/j.neuroimage.2018.10.023

Substance Abuse and Mental Health Services Administration. (2025). Key Substance Use and Mental Health Indicators in the United States: Results from the 2024 National Survey on Drug Use and Health. Center for Behavioral Health Statistics and Quality, Substance Abuse and Mental Health Services Administration. https://www.samhsa.gov/data/sites/default/files/reports/rpt56287/2024-nsduh-annual-national/2024-nsduh-annual-national-html-071425-edited/2024-nsduh-annual-national.htm?utm_source=chatgpt.com#alc1

Van Essen, D. C., Smith, S. M., Barch, D. M., Behrens, T. E. J., Yacoub, E., & Ugurbil, K. (2013). The WU-Minn Human Connectome Project: An overview. NeuroImage, 80, 62–79. 10.1016/j.neuroimage.2013.05.041

Wang, H.-T., Smallwood, J., Mourao-Miranda, J., Xia, C. H., Satterthwaite, T. D., Bassett, D. S., & Bzdok, D. (2020). Finding the needle in a high-dimensional haystack: Canonical correlation analysis for neuroscientists. NeuroImage, 216, 116745. 10.1016/j.neuroimage.2020.116745

Yeo, B. T. T., Krienen, F. M., Sepulcre, J., Sabuncu, M. R., Lashkari, D., Hollinshead, M., Roffman, J. L., Smoller, J. W., Zöllei, L., Polimeni, J. R., Fischl, B., Liu, H., & Buckner, R. L. (2011). The organization of the human cerebral cortex estimated by intrinsic functional connectivity. Journal of Neurophysiology, 106(3), 1125–1165. 10.1152/jn.00338.2011

