## Supplement for "Distinct Multimodal Brain Profiles Bridging Age of First Alcohol Use and Subsequent Substance Use in Young Adults"

**Methods**

**MRI data acquisition** was done using standardized HCP-YA imaging protocols on a Siemens 3T Skyra scanner to acquire high resolution structural T1-weighted images using a 3D MPRAGE acquisition with TE/TR/FA=2.14ms/2400ms/8°, 0.7 mm isotropic resolution and structural T2-weighted images using 3D T2-SPACE acquisition with TE/TR=565ms/3200ms, 0.7 mm isotropic resolution (Harms et al., 2018). Blood oxygenation level dependent (BOLD) functional MRI data were collected using a T2-weighted* gradient-echo echo-planar resting state MRI with multi-band acquisition with TE/TR/FA=33.1ms/720ms/52°, multi-band factor=8, 72 slices, 2mm isotropic resolution, with four 15-minute runs (2 with left-right phase encoding and 2 with right-left phase encoding) collected while participants were resting with eyes open with relaxed fixation for a total of one hour of resting state fMRI per individual (Glasser et al., 2013).

**Structural MRI** data underwent the 2025 HCP minimal preprocessing pipelines, including gradient distortion correction, motion correction, bias field correction, surface reconstruction, and multimodal surface registration. Cortical thickness and mid-thickness surface area measures were computed from the T1 images. Intracortical myelin was estimated using the T1w/T2w ratio (Glasser & Van Essen, 2011), all via FreeSurfer using the HCP structural processing framework. Additional processing details are available in the HCP 2025 release documentation (<https://humanconnectome.org/storage/app/media/documentation/s1200/HCP_S1200_Release_Reference_Manual.pdf>). Cortical thickness and surface area were selected due to their known associations with AFAU (Infante et al., 2018; Miller et al., 2024), while intracortical myelin is an understudied measure that undergoes important changes during brain development and may therefore be relevant to AFAU.

**Resting-state fMRI** data were obtained from the legacy HCP minimally preprocessed release, which included gradient distortion correction, motion correction, EPI distortion correction, surface-based registration, and projection to CIFTI grayordinates as described for the HCP minimal preprocessing pipeline in Glasser et al., 2013. One hour of resting state fMRI was collected per participant. After individual-level denoising with ICA+FIX and multi-modal cortical surface registration (MSMAll), group-ICA was performed using FSL’s MELODIC to estimate 50 components, and resulting group level maps were projected onto individuals’ data via standard dual regression (Nickerson et al., 2017) to generate individual-level spatial network maps for each run for each participant. The network maps from the four runs for each network of interest were then averaged together and fed into data fusion.

**Network Selection Rationale.** The ventromedial prefrontal is strongly associated with emotion and reward processing (Hiser & Koenigs, 2018) and includes Brodmann areas 9 and 10 which are linked to emotional integration and inhibition and memory integration and inhibition respectively (Ben Shalom & Skandalakis, 2025). The two resting state components we selected encompass nearly all of prefrontal cortex, except for Brodmann 8A which is associated with motor function (Ben Shalom & Skandalakis, 2025). As shown in Supplemental Figure 2, our ventromedial DMN component strongly overlaps with Default C of the Yeo-17 parcellation (Yeo et al., 2011) and includes much of medial/ventromedial prefrontal cortex, while our frontal SN component strongly overlaps with the Salience B network of Yeo-17, which is a sub-network of the SN that includes lateral prefrontal areas. As such, while we did not include frontoparietal networks in our data fusion, the individual level DMN and SN maps from dual regression will reflect both within-network connectivity and DMN and SN connectivity with other regions of the brain (including those in the cognitive control networks).

**Train, test, external validation sets.** Recruitment focused on twins and siblings, resulting in extensive family structure within the HCP-YA. To avoid information leakage arising from family relatedness, participants were divided into 60% training, 20% testing, and 20% external validation datasets, with family members assigned to the same data split. Family structure was accounted for with Block permutations using the procedure described by Winkler et al. 2015, implemented in Python with a function adapted from the permuteHCP.py code (<https://github.com/tyo8/brain_representations/blob/main/src_py/calculate/permuteHCP.py>) and modified for the present study. Splits were also balanced on age and sex, with no significant differences observed across samples.

**Supplemental Figure 1.** The two resting state ICA components used in SBF analysis overlaid onto the MSMAll parcellation. As mentioned above, Component 12 most strongly overlaps with Yeo-17 DMN-A and Component 27 with Yeo-17 SN-B.

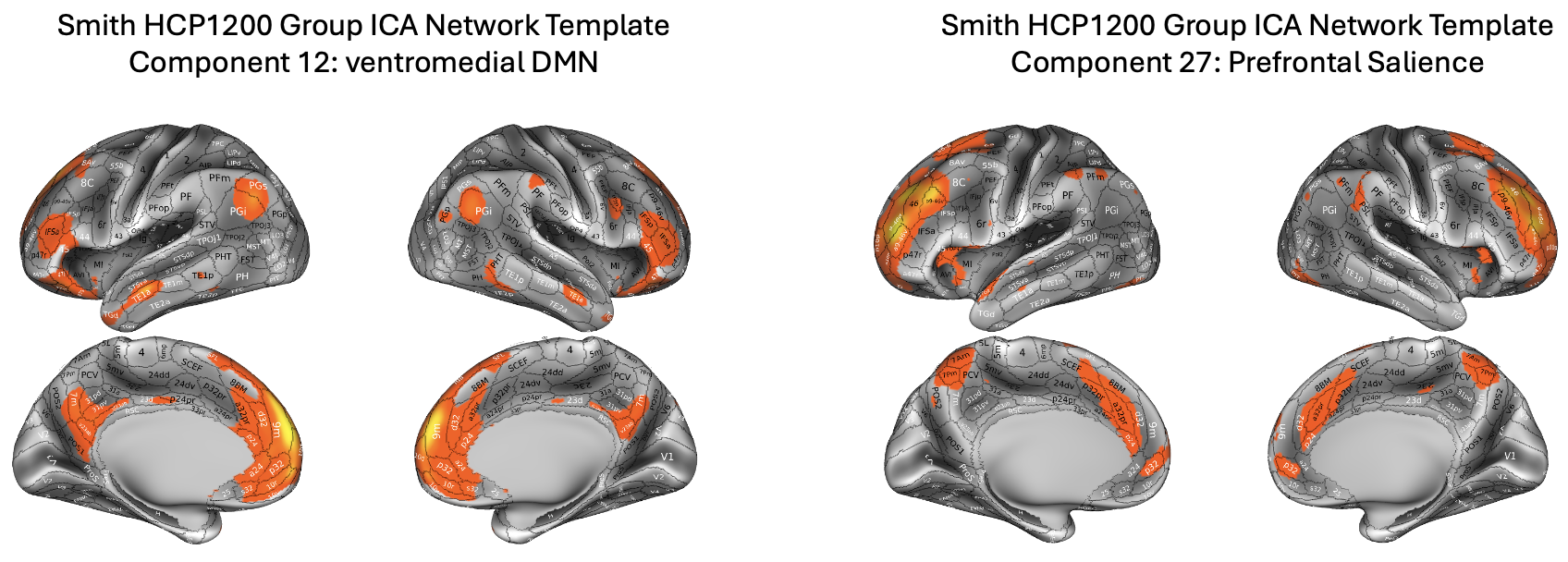

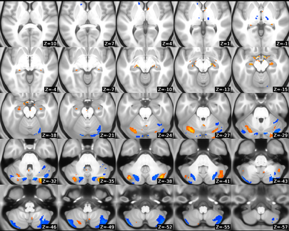

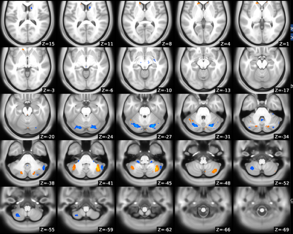

**Supplemental Figure 2.** Dice overlap of the Smith d50 ICA components with Yeo-17.

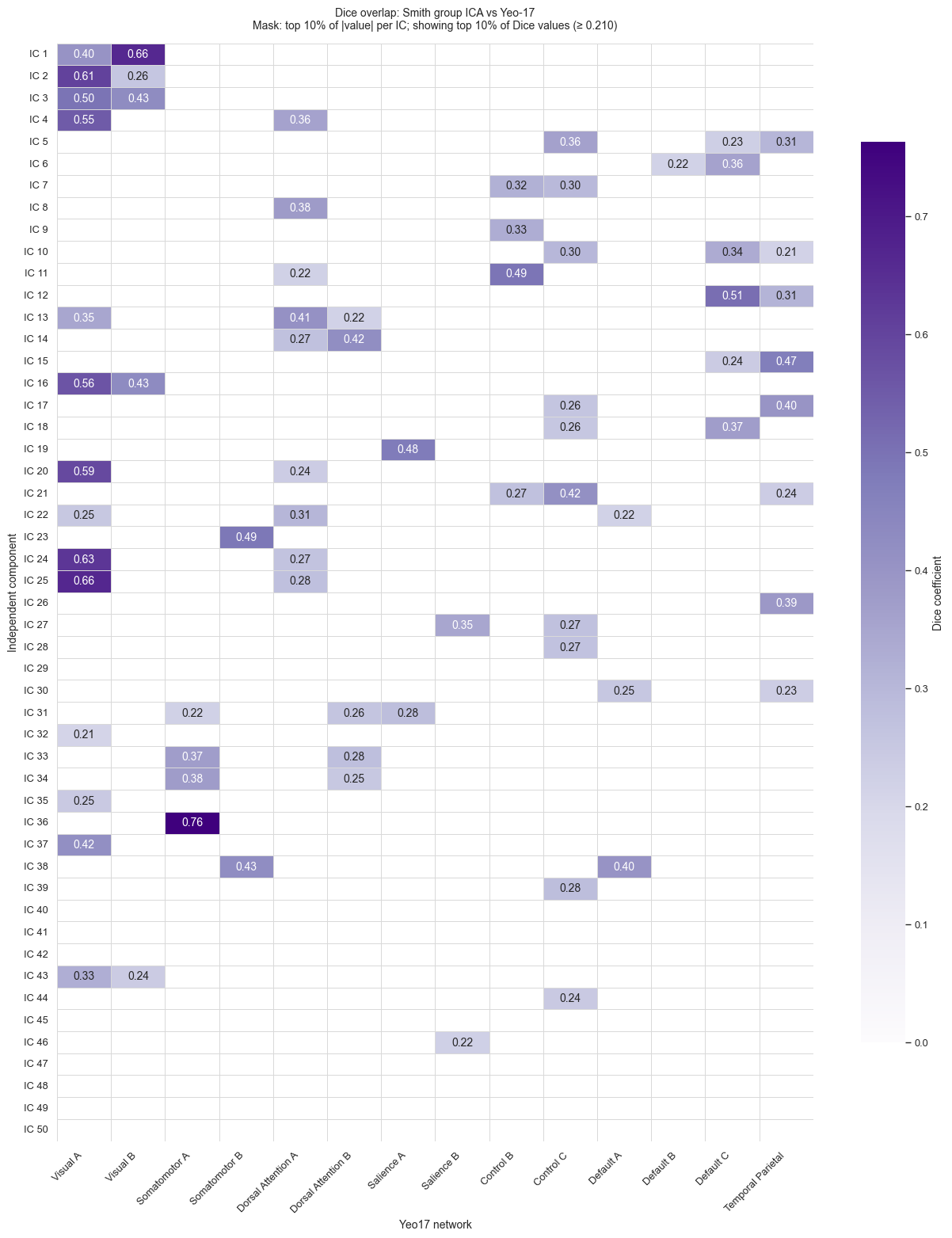

**Supplemental Figure 3.** SBF component loadings for each participant plotted against their associated SHAP values, for the top 5 components.

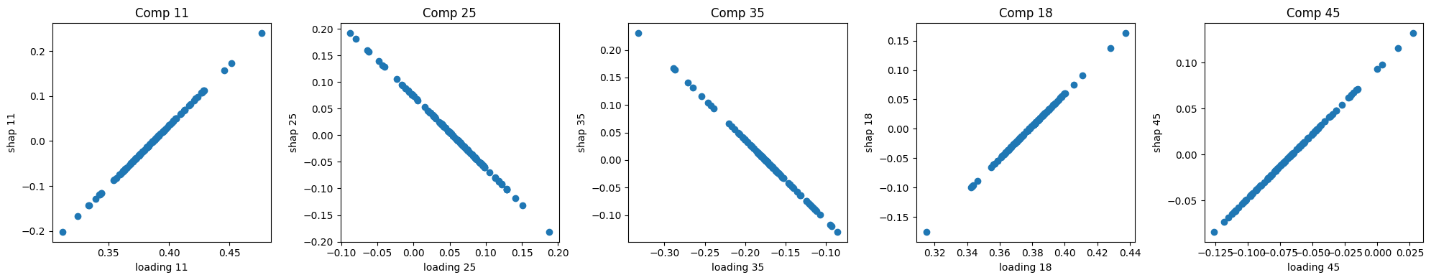

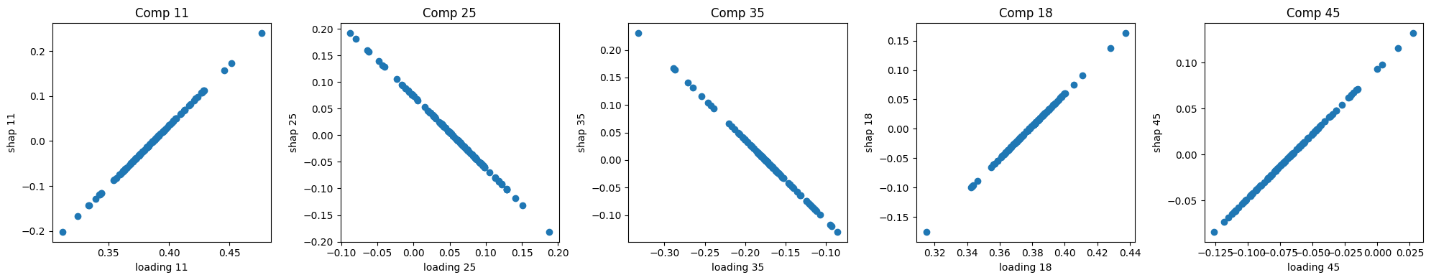

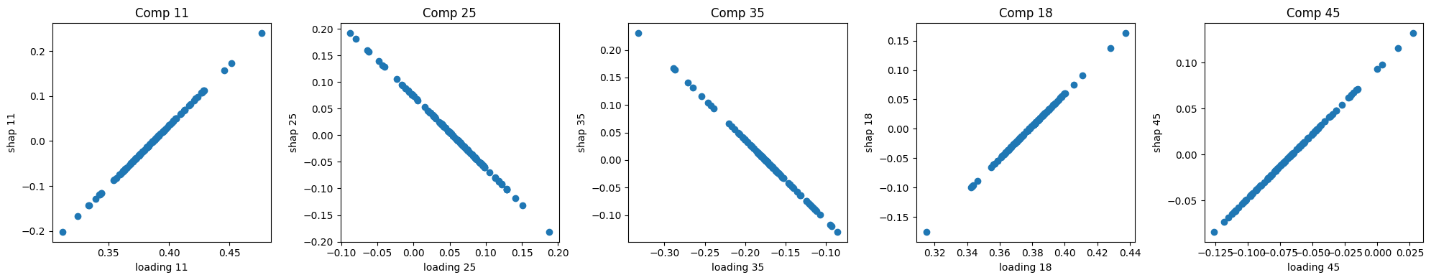

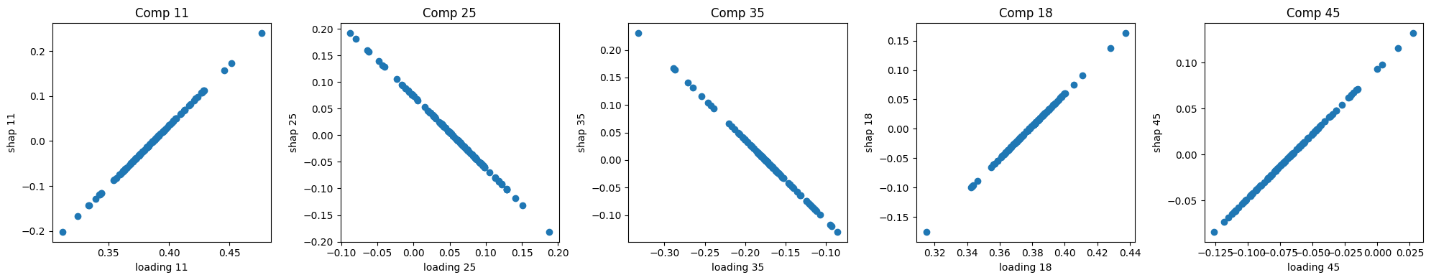

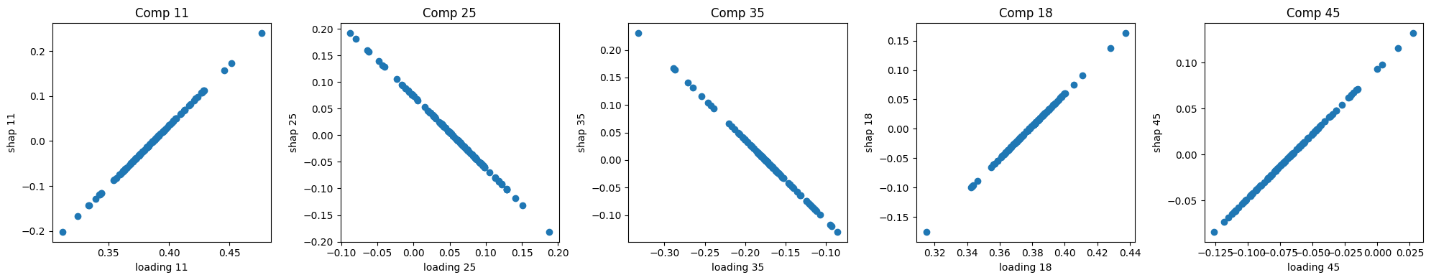

**Supplemental Figure 4.** Top 5 SBF components with Glasser parcellation underlay. Note that cortical thickness, mid-thickness surface area and intracortical myelin are cortex only modalities, whereas DMN and SN networks include sub-cortical areas (Supp Fig 1 above). However, none of the SBF components involving DMN and SN showed sub-cortical effects so these areas are not shown in the component maps below.

**Brain Component 11**

Cortical Thickness (0.48)

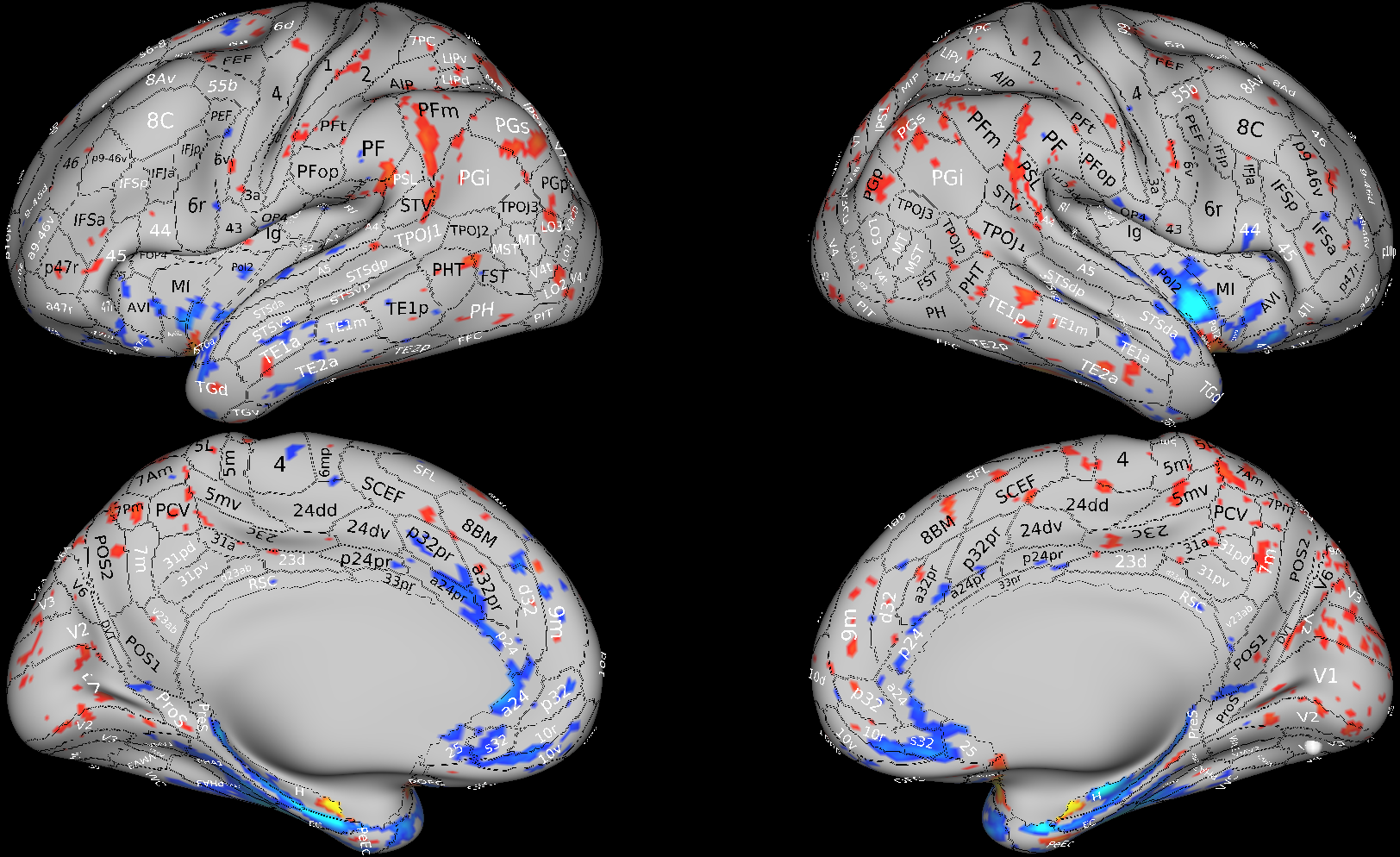

Surface Area (0.26)

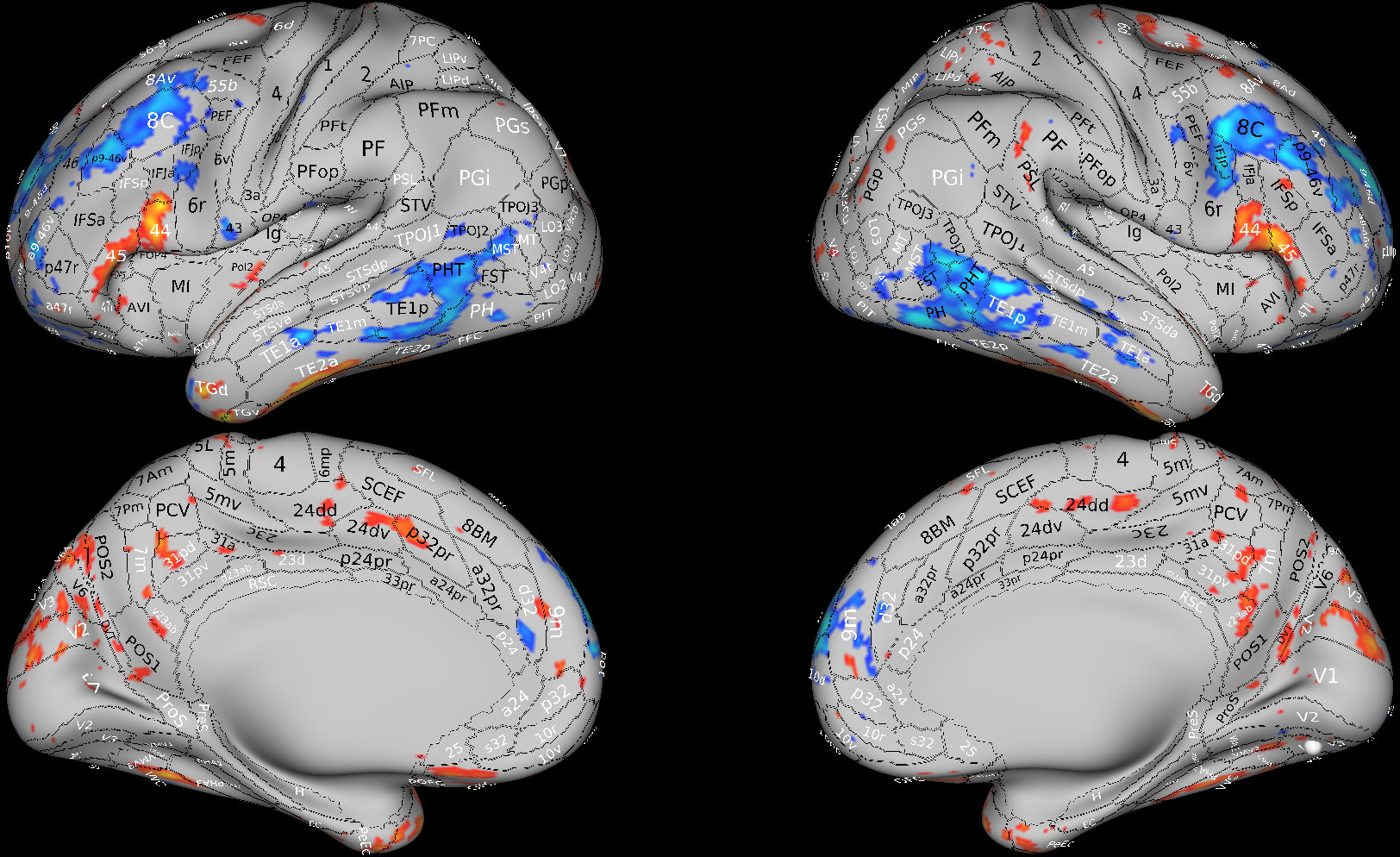

Salience Network (0.26)

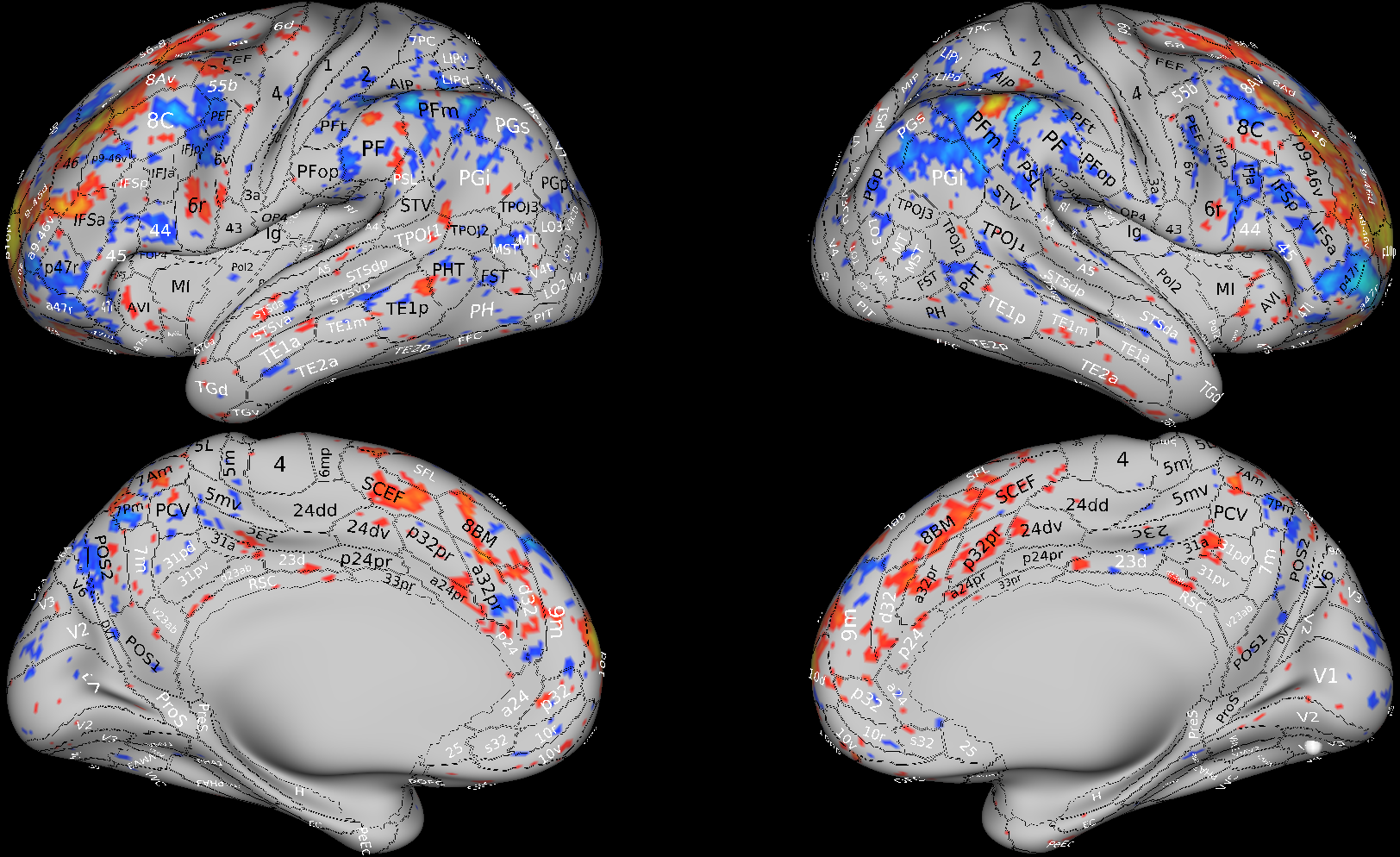

**Brain Component 25**

Surface area (1.0)

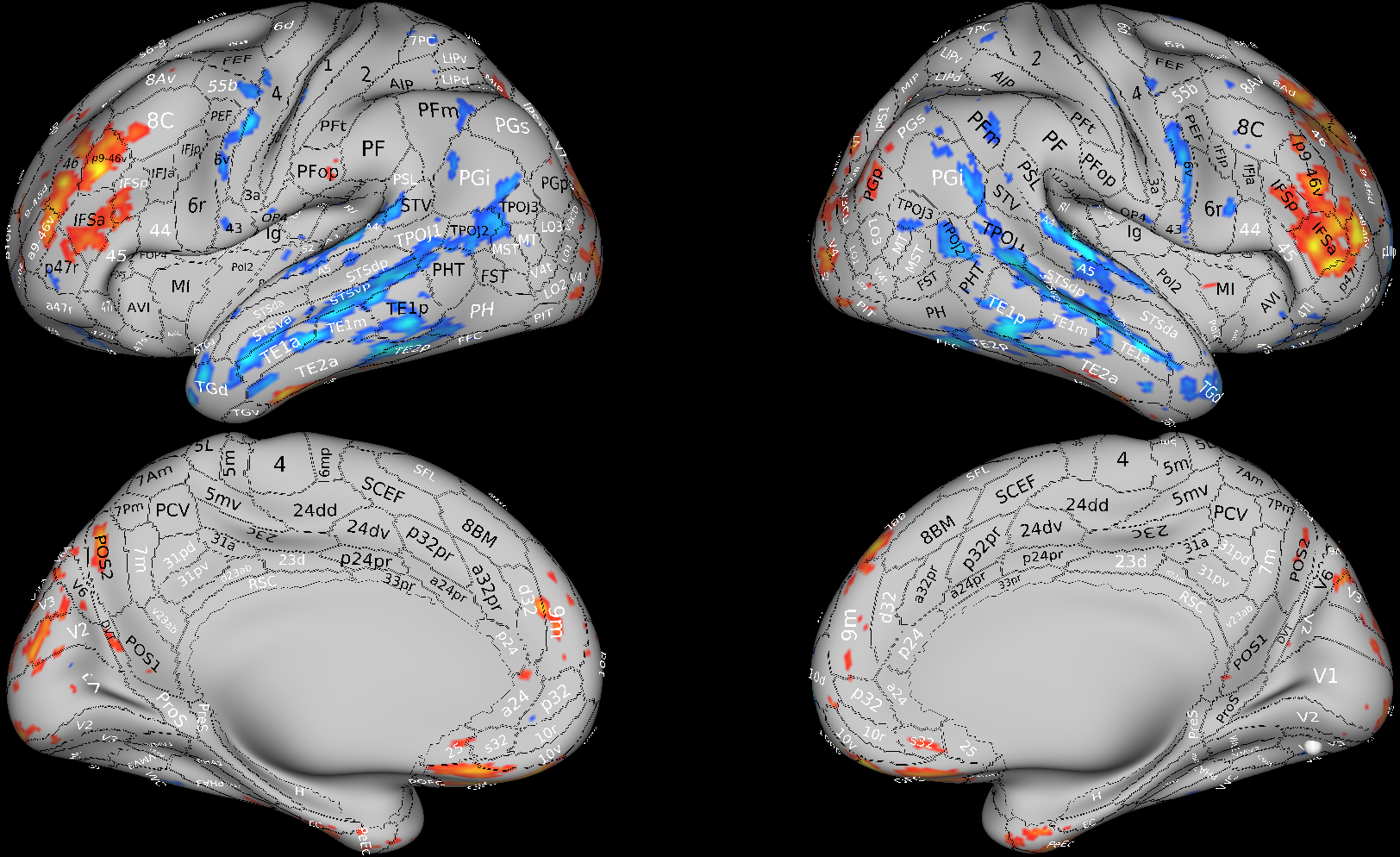

**Brain Component 35**

Default Mode Network (0.94)

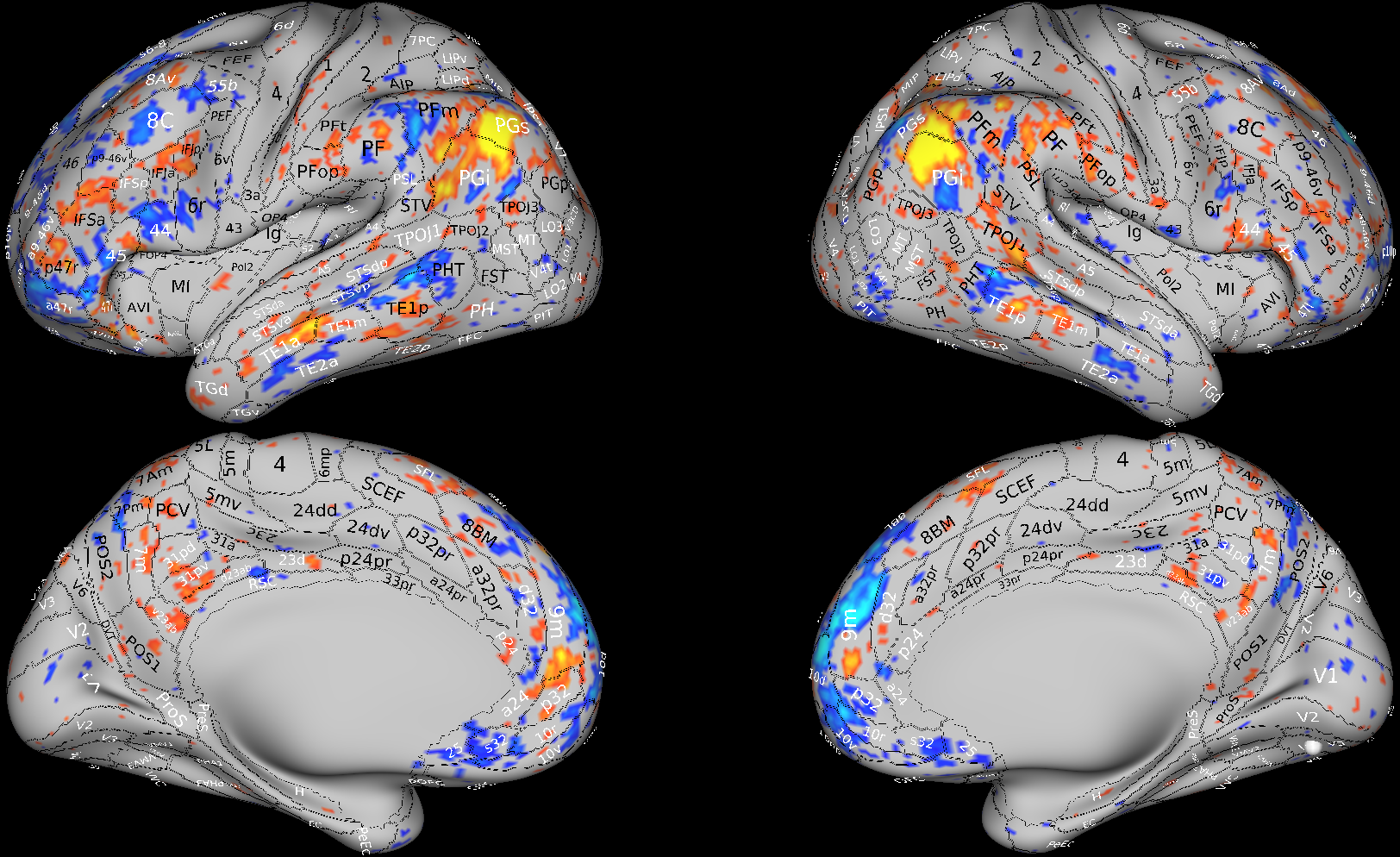

**Brain Component 18**

Intracortical Myelin (0.88)

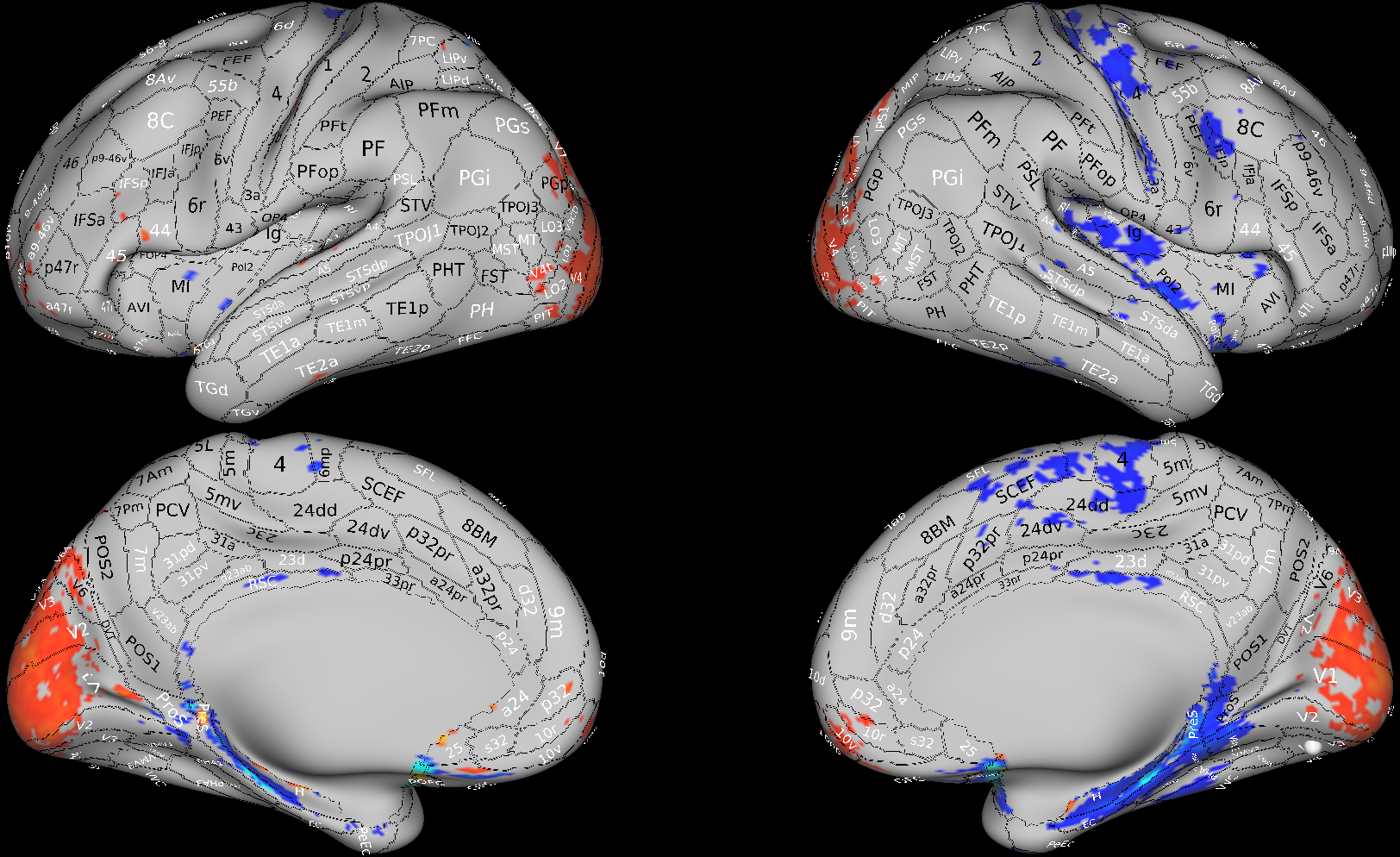

**Brain Component 45**

Salience Network (0.60)

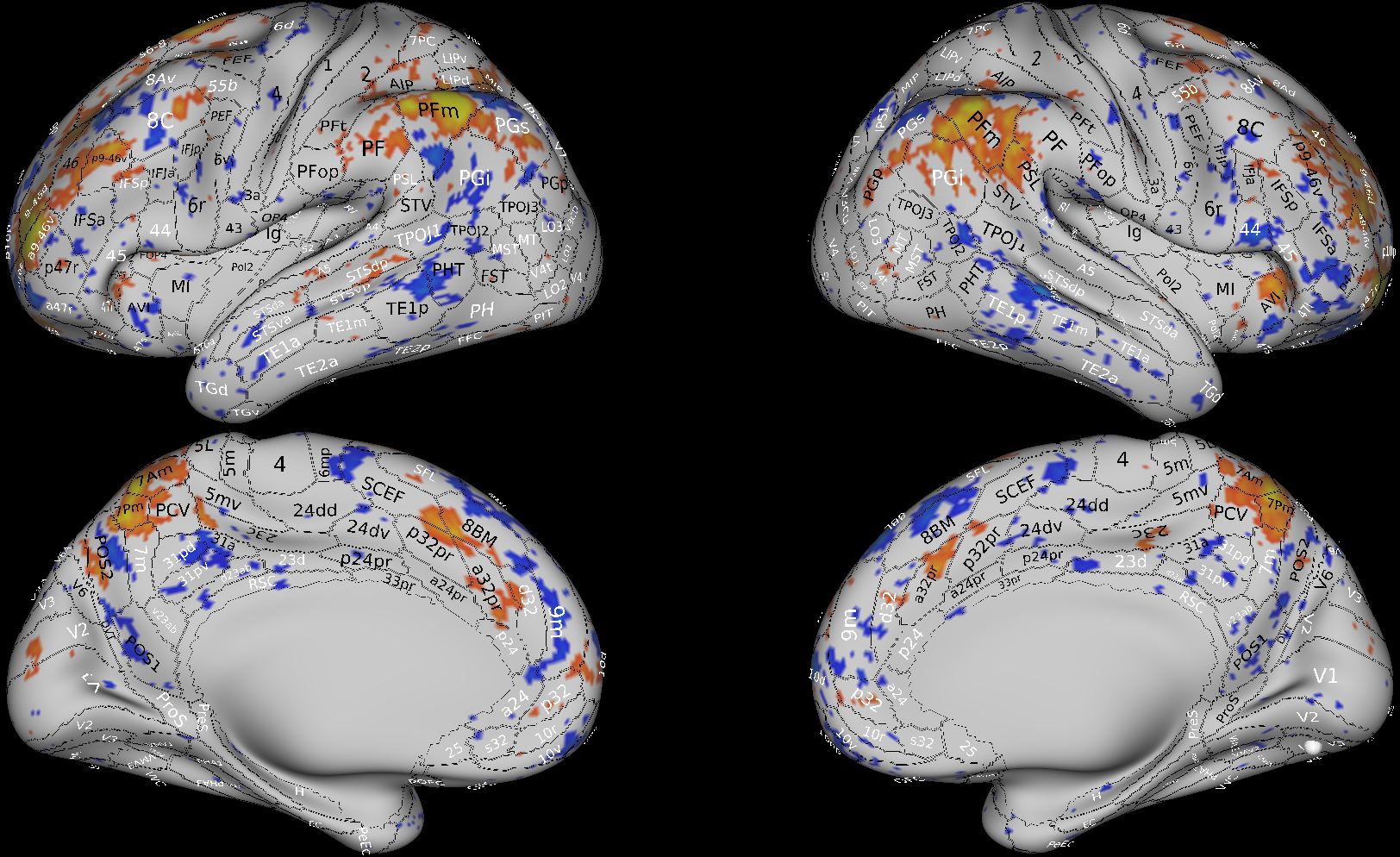

Surface Area (0.22)

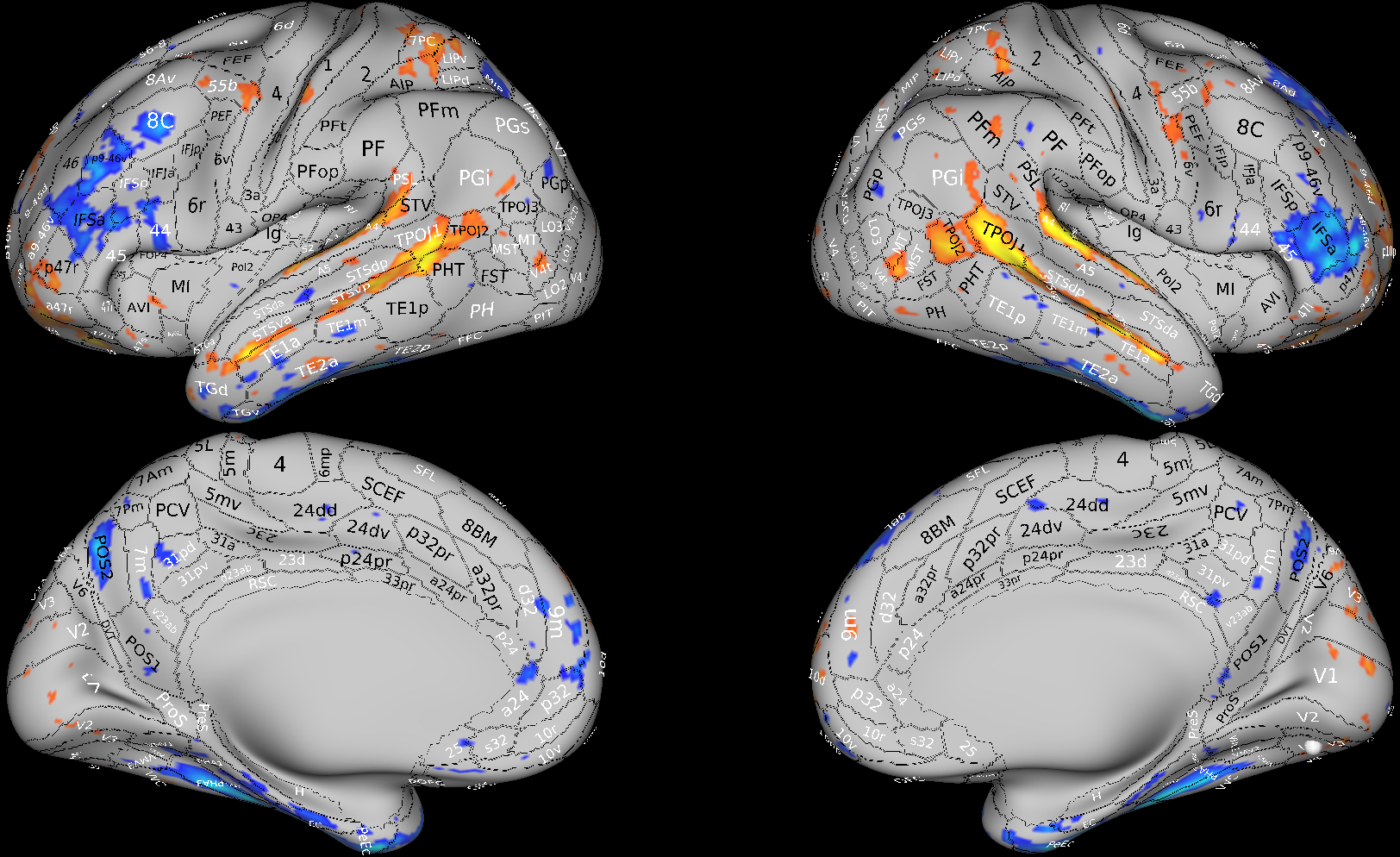

Default Mode Network (0.10)

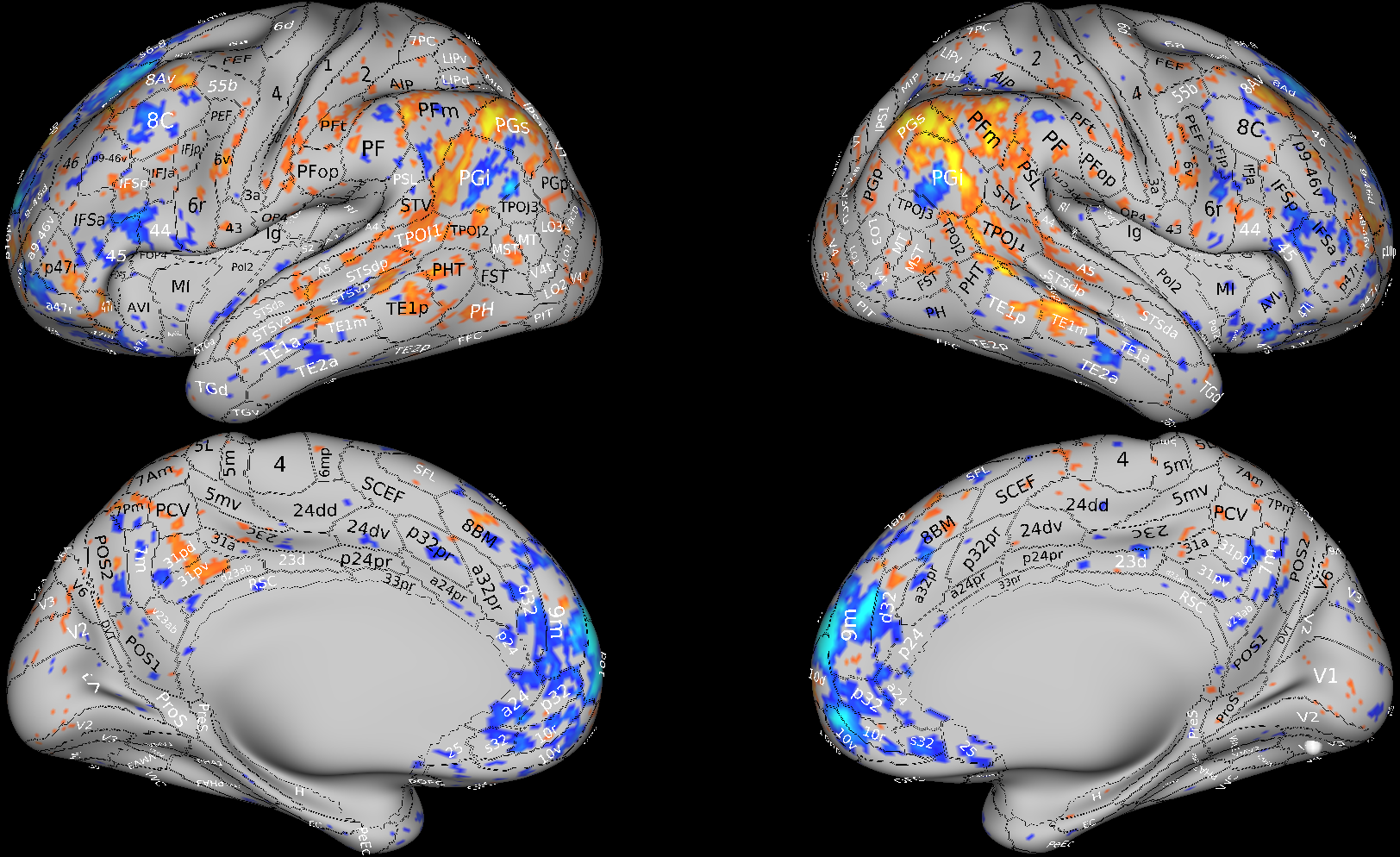

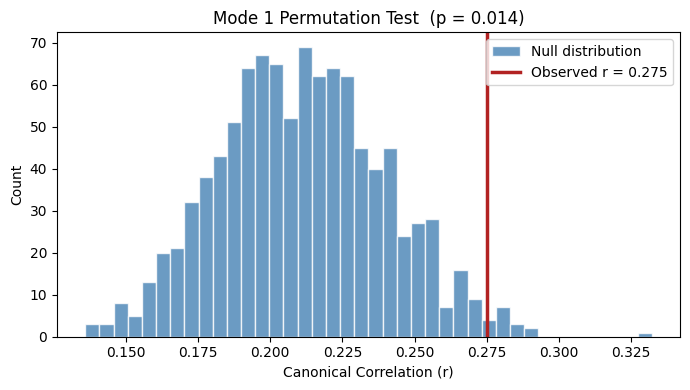

**Supplemental Figure 5.** Permutation tests for the 3 significant CCA modes.

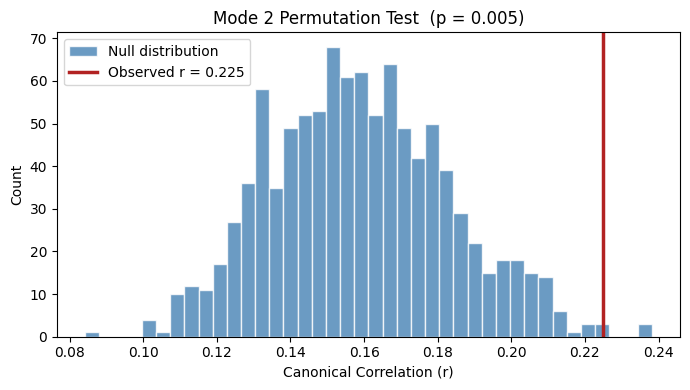

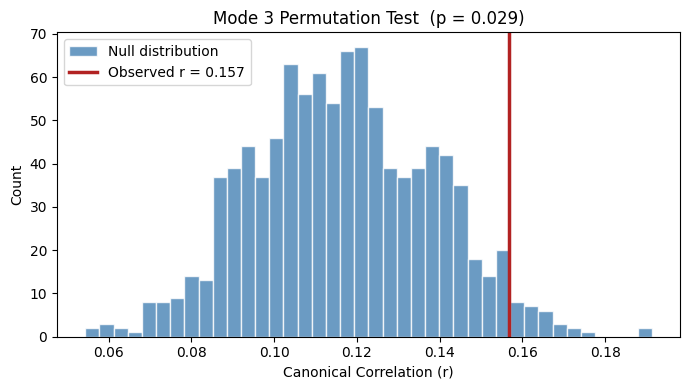
**Supplemental Figure 6.** Unthresholded loadings of the 3 significant CCA modes.

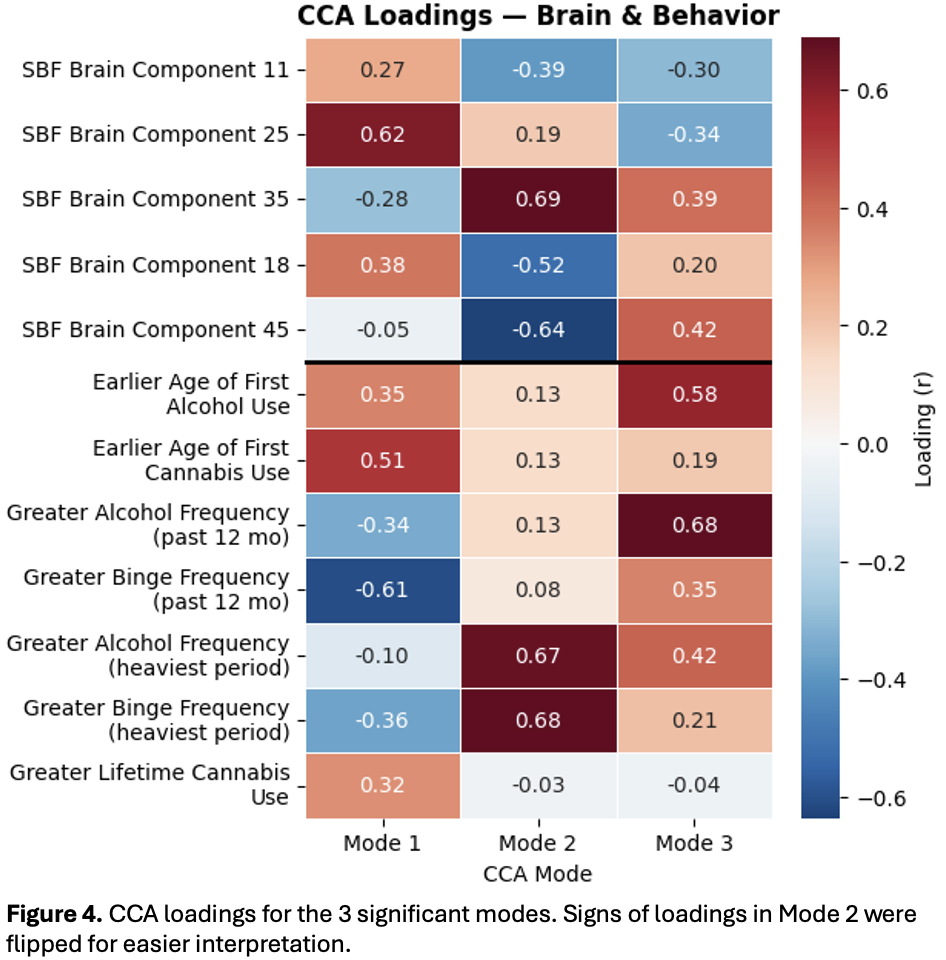
